# Pharmacist-Led Comprehensive Medication Management and Patient Outcomes in Pediatric Intensive Care Unit Patients: A Multicenter Cohort Study

**DOI:** 10.64898/2026.10.21.26366147

**Authors:** Kristine A. Parbuoni, Kelli R. Henry, Mojdeh S. Heavner, Andrea Sikora, Zhetao Chen, Xianyan Chen, Huong T. Duong, Elizabeth Farrington, Amy Kiskaddon, Jennifer Majchrzak, OPTIM Investigator Team, Susan E. Smith

**Affiliations:** University of Maryland School of Pharmacy, Department of Practice, Sciences, and Health Outcomes Research, Baltimore, MD, USA; University of Colorado School of Medicine, Department of Biomedical Informatics, Aurora, CO, USA; University of Georgia College of Public Health, Department of Epidemiology & Biostatistics, Athens, GA, USA; University of Georgia College of Public Health, Department of Epidemiology & Biostatistics, Augusta, GA, USA; University of Georgia College of Pharmacy, Department of Clinical and Administrative Pharmacy, Athens, GA, USA; Novant Health New Hanover Regional Medical Center, Wilmington, NC, USA; Johns Hopkins University School of Medicine, Baltimore, MD, USA Johns Hopkins All Children’s Hospital, St. Petersburg, FL, USA; University of Maryland Upper Chesapeake Health, Bel Air, MD, USA

**Keywords:** clinical pharmacists, PICU, NICU, patient outcomes, comprehensive medication management

## Abstract

**Objective:** Clinical pharmacists improve medication management and safety in intensive care units, yet their role in neonatal and pediatric intensive care remains highly variable across institutions. Evidence supporting standardized clinical pharmacy practice models in these settings is limited. This study evaluated the association between pharmacist service coverage and clinical outcomes in critically ill pediatric patients.

**Design:** Multicenter, observational study with prospective and retrospective data collection.

**Setting:** ICUs at participating institutions in the United States and internationally.

**Patients:** Patients <18 years of age

**Intervention:** None

**Measurements:** The primary exposure was pharmacist-led comprehensive medication management (CMM) with daily participation in multidisciplinary rounds. The secondary exposure was the pharmacist-to-ICU-patient ratio. The primary outcome was hazard of discharge alive (HDA) from the ICU. Secondary outcomes included in-hospital mortality, hospital HDA, and hazard of extubation alive (HEA). Fine-Gray competing risks models and multivariable Firth logistic regression were used for adjusted analyses.

**Main results:** A total of 1,459 patients cared for by 69 clinical pharmacists across 31 institutions were included. Daily CMM with rounding was associated with a higher HDA from the ICU (HR 2.60, 95% CI 2.17-3.10; p<0.001), higher HDA from the hospital (HR 2.25, 95% CI 1.83-2.75; p<0.001), and a higher HEA (HR 1.21, 95% CI 1.10-1.33; p<0.001), but not with in-hospital mortality (OR 0.44, 95% CI 0.14-1.36; p=0.154). The pharmacist-to-ICU-patient ratio was not significantly associated with ICU HDA (HR 1.00, 95% CI 0.99-1.01; p=0.88) or any secondary outcome.

**Conclusions:** Daily pharmacist-provided CMM with multidisciplinary ICU rounding was associated with improved discharge outcomes among critically ill pediatric and neonatal patients, whereas pharmacist-to-ICU-patient ratio alone was not. These findings support standardized integration of clinical pharmacists into daily multidisciplinary rounds in this setting.

## INTRODUCTION

“Every critically ill patient admitted to a critical care service requires the care of a critical care pharmacist.” This consensus recommendation statement, endorsed by the American Association of Critical-Care Nurses (AACN), American College of Clinical Pharmacy (ACCP), American Society of Health-System Pharmacists (ASHP), Institute for Safe Medication Practice (ISMP), and Society of Critical Care Medicine (SCCM)^1^, highlights the essential role of clinical pharmacists in the care of critically ill patients. While much of the evidence informing this statement is derived from adult populations, neonatal and pediatric patients have unique medication-related risks that warrant equal attention.^2,3^ Despite their demonstrated value in medication management and safety, clinical pharmacists frequently face pressure to justify their role.^4^ This challenge may be especially pronounced in the neonatal intensive care unit (NICU) and pediatric intensive care unit (PICU), where clinical pharmacist practice models vary across institutions and limited standardized approaches complicate workload allocation between direct and indirect patient care responsibilities.^5^

Medication management in children is inherently demanding, as most medications lack an FDA-approved pediatric indication, resulting in exposure to an average of 4.5 off-label medications among pediatric critical care patients.^6,7^ These patients also face nearly three times the medication error risks as adults, owing to limited pediatric pharmacokinetic data, complex weight-based dosing, and adult-oriented medication formulations.^8,9^ These factors emphasize the need for consistent clinical pharmacy services, yet limited pediatric-specific evidence has hindered efforts to justify and optimize NICU/PICU care. Comprehensive medication management (CMM),^10–12^ a clinical pharmacy service in which each patient’s medications are individually assessed for appropriateness, effectiveness, safety, and ability to be taken as intended, has become the standard of adult ICU care.^1,13–15^ Nonetheless, evidence supporting this service in neonatal and pediatric populations remains limited. Similarly, despite the proven benefits of multiprofessional care on adult ICU outcomes, its impact in non-adult populations remain poorly studied.^16–22^ The purpose of this study is to examine the relationship between pharmacist coverage, including daily CMM with interdisciplinary rounds participation, and patient outcomes in the NICU and PICU. The ultimate goal is to inform pediatric pharmacy practice standards and guide workload optimization to ensure delivery of the highest quality care.

## MATERIALS AND METHODS

This study represents the pediatric cohort of The Optimizing Pharmacist Team-Integration for ICU Patient Management (OPTIM) study.^23^ OPTIM was a multicenter, observational cohort study, using both prospective and retrospective data collection strategies to evaluate the relationship between clinical pharmacist staffing practices and ICU patient-centered outcomes. A detailed protocol was previously published with an analysis of adult patients.^23,24^ The University of Georgia (UGA) Institutional Review Board (IRB) determined this study to be exempt. All participating institutions completed a data use agreement and IRB per institutional requirements.

Data were collected and managed using REDCap (Research Electronic Data Capture) tools hosted at the UGA.^25^ Participants were recruited through targeted outreach to postgraduate year two (PGY2) critical care pharmacy residency program directors and professional organization members including the ACCP Critical Care Practice and Research Network and the SCCM Clinical Pharmacy and Pharmacology Section. Clinical pharmacists whose primary responsibility was providing pharmaceutical care for ICU patients and who were not enrolled in a training program were eligible to participate. This study included ICU patients who were less than 18 years old with ICU stay of at least 24 hours, excluding those transitioned to comfort measures within the first 24 hours of ICU care.

Data were collected in three phases:

### Phase 1

A series of three surveys captured institution characteristics including pharmacy staffing models, ICU team characteristics, and pharmacist characteristics.

### Phase 2

For 100 consecutive days between August 2023 and August 2024, each participating pharmacist recorded observational workload and patient data. The pharmacist collected both patient data on all eligible patients and pharmacist staffing data on each working day. Patient data included the patient-specific pharmacist-to-patient ratio, whether the patient received CMM, the nurse-to-patient ratio, and the composition of the medical prescribing team. Pharmacist staffing data included the number of interprofessional rounding teams assigned to the pharmacist, the number of ICU and non-ICU patients assigned, and other relevant workload indicators. If a participating pharmacist was not working on a given day, they retrospectively recorded the working pharmacist’s workload variables on their next workday.

### Phase 3

Retrospective data from the electronic health record (EHR) for each study patient were collected through January 2025 and included baseline demographics, severity of illness using the Pediatric Risk of Mortality III (PRISM III) score, medication regimen complexity-intensive care unit (MRC-ICU) score during the first 24 hours of the ICU admission, and the defined outcome data.^26,27^

The primary outcome of this study was hazard of discharge alive (HDA) from the ICU. Because in-hospital death precludes discharge alive, HDA provided a more appropriate assessment of hospital stay than conventional length-of-stay (LOS) analyses when mortality differs between groups. Secondary outcomes included in-hospital mortality, hospital HDA, and hazard of extubation alive (HEA).

The primary variable was pharmacist-provided CMM, including daily attendance on multidisciplinary rounds every day of ICU stay (daily CMM with rounding). The secondary variable was patient-specific pharmacist-to-ICU-patient ratio (PPR) averaged over the ICU stay. Covariates included patient demographics, illness severity, institutional characteristics, and pharmacy and nursing staffing. Detailed covariates can be found in **Table 3**.

Fine-Gray Competing Risks Models were used to examine factors associated with the HDA from the ICU and hospital and the HEA, with mortality treated as a competing event for both outcomes. Multivariable Firth logistic regression was used to evaluate factors associated with in-hospital mortality. Firth logistic regression was selected over conventional logistic regression due to the expected rarity of mortality events, reducing small-sample bias and the risk of unstable or infinite estimates from quasi-complete separation. Results are reported as subdistribution hazard ratios (sHRs) with 95% confidence intervals. Missing covariate data were handled using multiple imputation by chained equations, generating 10 imputed datasets. Outcome variables were not imputed. Each analysis model was fitted separately within each imputed dataset, and coefficient estimates and variances were combined using Rubin’s rules. Both primary and subgroup analyses used multiple imputation. All analyses were performed using R version 4.4.1 with mice (multivariate imputation), riskRegression and cmprsk (competing risks), logistf (Firth’s penalized logistic regression), and mitools (multiply imputed data manipulation) packages. A two-side p-value less than 0.05 was considered statistically significant.

PICU and NICU patients underwent subgroup analysis using multiple imputed datasets **(Supplementary Appendix)**, including demographics, missing data, regression models for mortality, and Fine-Gray competing-risks models for ICU HDA, hospital HDA, and HEA.

## RESULTS

The OPTIM dataset included a total of 33,464 patients at 64 institutions. After selecting for eligible pediatric patients, 1,459 pediatric patients cared for by 69 clinical pharmacists across 31 institutions were included **(Figure 1).** Of these institutions, 30 were located in the US, representing the majority of geographic regions except non-continental regions. The international site included was in Saudi Arabia. Most sites were academic medical centers (67.7%), followed by community teaching hospitals (22.6%). More than half had over 600 hospital beds (58.1%) and over 100 ICU beds (61.3%). In terms of critical care pharmacy practice model, 83.9% of institutions conducted bedside rounds 5 days per week on day shift. Weekend clinical coverage varied, with the most common coverage model consisting of pharmacokinetic monitoring and prospective medication order verification (41.9%). Details on institution characteristics are presented in **Table 1**.

**Figure 1.**
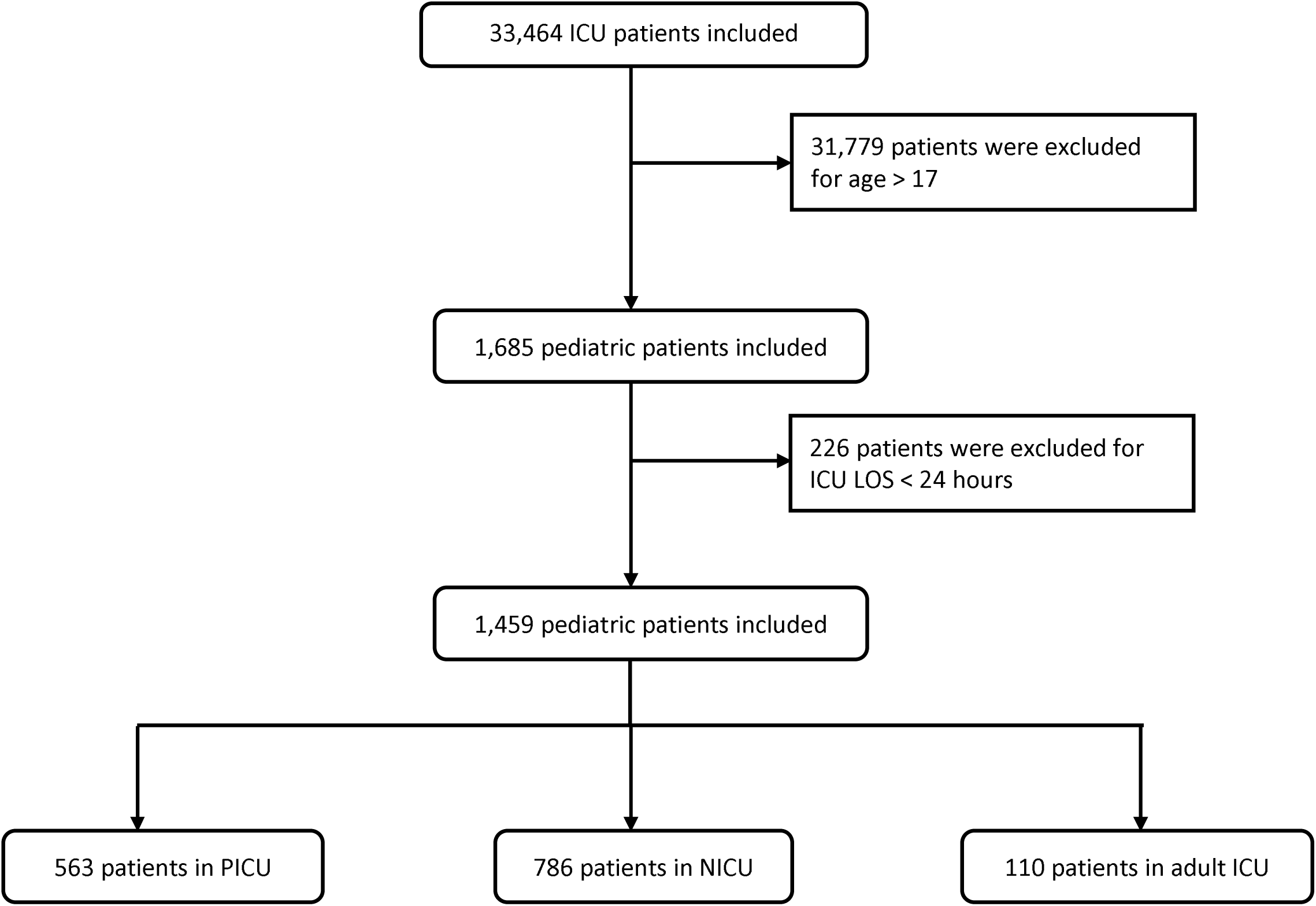
CONSORT Diagram. A total of 1,459 patients cared for by 69 clinical pharmacists across 31 institutions were included in the study. Of those, 786 were admitted to pediatric intensive care units (ICUs) and 563 to the neonatal ICUs and were included in the subgroup analyses. The remaining 110 patients were pediatric patients admitted to adult ICUs, with the majority from an adult trauma ICU. ICU: intensive care unit, LOS: length of stay, NICU: neonatal intensive care, PICU: pediatric intensive care unit

**Table 1.**
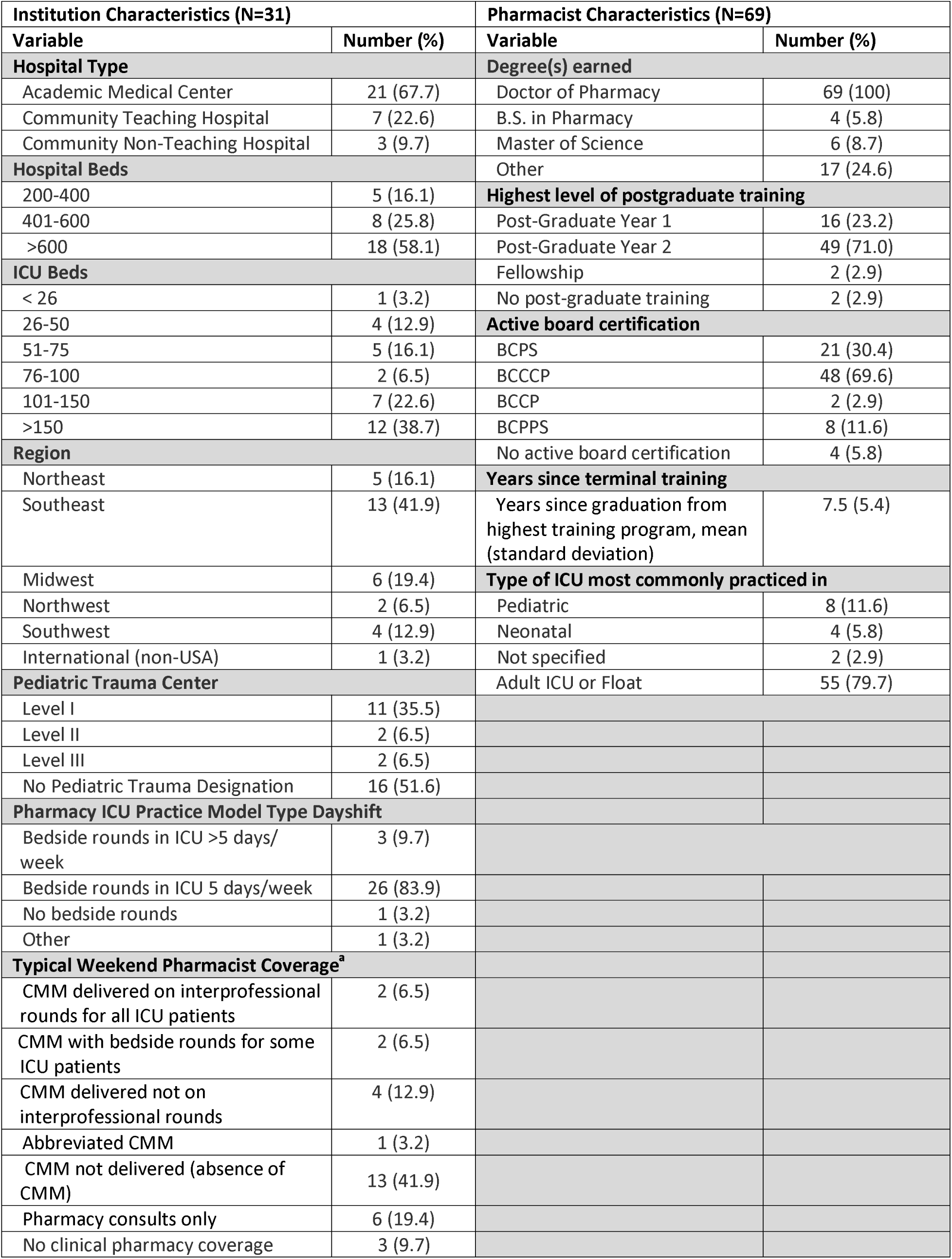
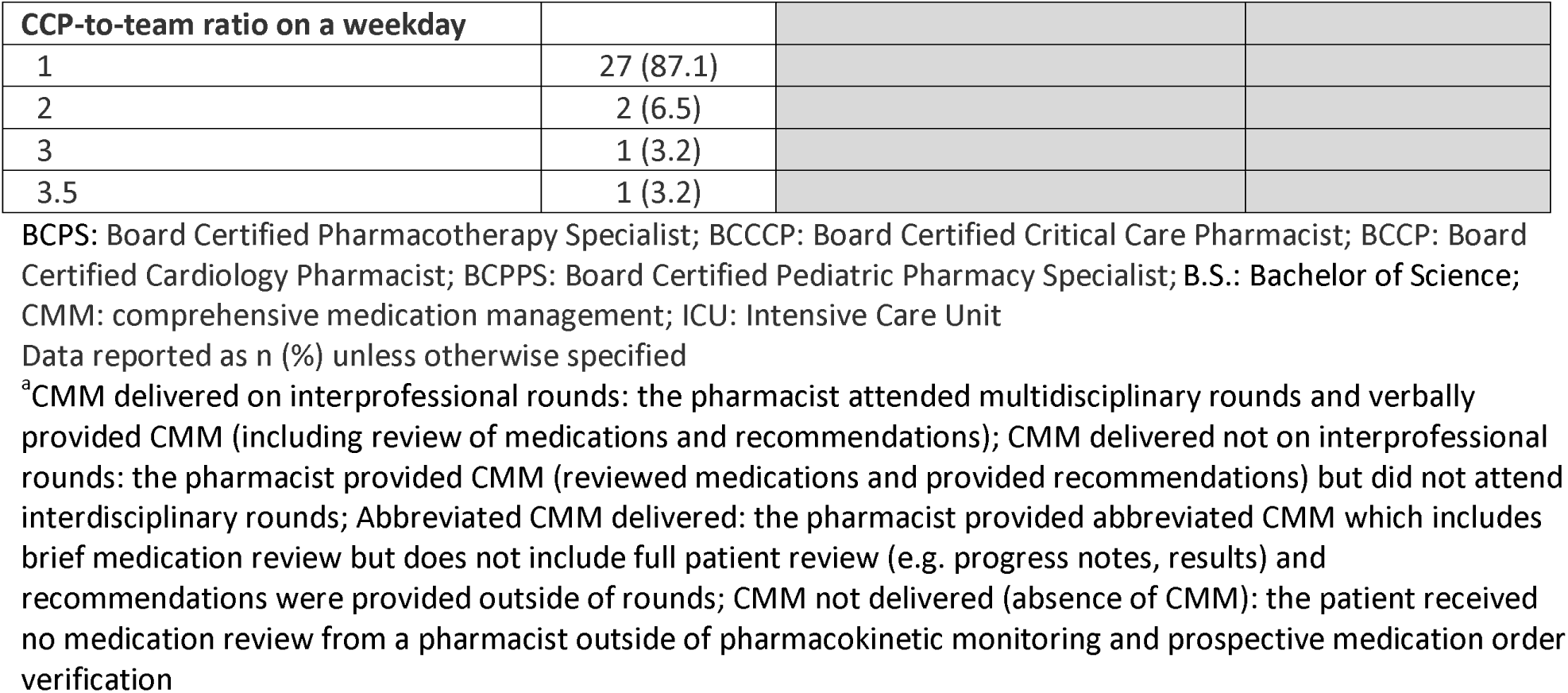
Institution and Pharmacist Characteristics.

| Institution Characteristics (N=31) |  | Pharmacist Characteristics (N=69) |  |
| --- | --- | --- | --- |
| Variable | Number (%) | Variable | Number (%) |
| **Hospital Type** |  | **Degree(s) earned** |  |
| Academic Medical Center | 21 (67.7) | Doctor of Pharmacy | 69 (100) |
| Community Teaching Hospital | 7 (22.6) | B.S. in Pharmacy | 4 (5.8) |
| Community Non-Teaching Hospital | 3 (9.7) | Master of Science | 6 (8.7) |
| **Hospital Beds** |  | Other | 17 (24.6) |
| 200-400 | 5 (16.1) | **Highest level of postgraduate training** |  |
| 401-600 | 8 (25.8) | Post-Graduate Year 1 | 16 (23.2) |
| >600 | 18 (58.1) | Post-Graduate Year 2 | 49 (71.0) |
| **ICU Beds** |  | Fellowship | 2 (2.9) |
| < 26 | 1 (3.2) | No post-graduate training | 2 (2.9) |
| 26-50 | 4 (12.9) | **Active board certification** |  |
| 51-75 | 5 (16.1) | BCPS | 21 (30.4) |
| 76-100 | 2 (6.5) | BCCCP | 48 (69.6) |
| 101-150 | 7 (22.6) | BCCP | 2 (2.9) |
| >150 | 12 (38.7) | BCPPS | 8 (11.6) |
| **Region** |  | No active board certification | 4 (5.8) |
| Northeast | 5 (16.1) | **Years since terminal training** |  |
| Southeast | 13 (41.9) | Years since graduation from highest training program, mean (standard deviation) | 7.5 (5.4) |
| Midwest | 6 (19.4) | **Type of ICU most commonly practiced in** |  |
| Northwest | 2 (6.5) | Pediatric | 8 (11.6) |
| Southwest | 4 (12.9) | Neonatal | 4 (5.8) |
| International (non-USA) | 1 (3.2) | Not specified | 2 (2.9) |
| **Pediatric Trauma Center** |  | Adult ICU or Float | 55 (79.7) |
| Level I | 11 (35.5) |  |  |
| Level II | 2 (6.5) |  |  |
| Level III | 2 (6.5) |  |  |
| No Pediatric Trauma Designation | 16 (51.6) |  |  |
| **Pharmacy ICU Practice Model Type Dayshift** |  |  |  |
| Bedside rounds in ICU >5 days/week | 3 (9.7) |  |  |
| Bedside rounds in ICU 5 days/week | 26 (83.9) |  |  |
| No bedside rounds | 1 (3.2) |  |  |
| Other | 1 (3.2) |  |  |
| **Typical Weekend Pharmacist Coverage<sup>a</sup>** |  |  |  |
| CMM delivered on interprofessional rounds for all ICU patients | 2 (6.5) |  |  |
| CMM with bedside rounds for some ICU patients | 2 (6.5) |  |  |
| CMM delivered not on interprofessional rounds | 4 (12.9) |  |  |
| Abbreviated CMM | 1 (3.2) |  |  |
| CMM not delivered (absence of CMM) | 13 (41.9) |  |  |
| Pharmacy consults only | 6 (19.4) |  |  |
| No clinical pharmacy coverage | 3 (9.7) |  |  |

| CCP-to-team ratio on a weekday |  |
| --- | --- |
| 1 | 27 (87.1) |
| 2 | 2 (6.5) |
| 3 | 1 (3.2) |
| 3.5 | 1 (3.2) |

BCPS: Board Certified Pharmacotherapy Specialist; BCCCP: Board Certified Critical Care Pharmacist; BCCP: Board Certified Cardiology Pharmacist; BCPPS: Board Certified Pediatric Pharmacy Specialist; B.S.: Bachelor of Science; CMM: comprehensive medication management; ICU: Intensive Care Unit
Data reported as n (%) unless otherwise specified
<sup>a</sup>CMM delivered on interprofessional rounds: the pharmacist attended multidisciplinary rounds and verbally provided CMM (including review of medications and recommendations); CMM delivered not on interprofessional rounds: the pharmacist provided CMM (reviewed medications and provided recommendations) but did not attend interdisciplinary rounds; Abbreviated CMM delivered: the pharmacist provided abbreviated CMM which includes brief medication review but does not include full patient review (e.g. progress notes, results) and recommendations were provided outside of rounds; CMM not delivered (absence of CMM): the patient received no medication review from a pharmacist outside of pharmacokinetic monitoring and prospective medication order verification

**Figure 2.**
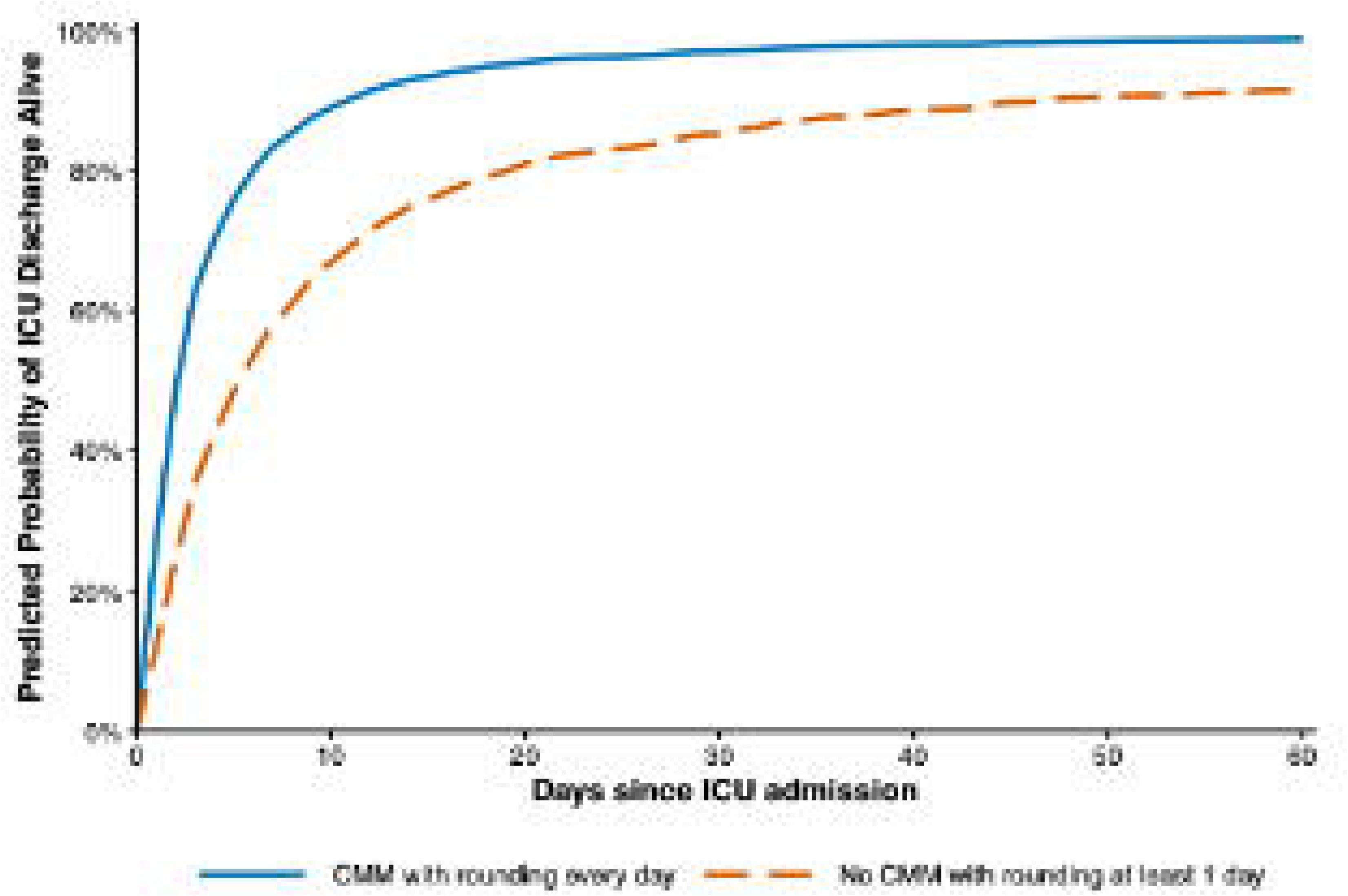
Cumulative Incidence Function (CIF) Predicted Probability of Discharge Alive from ICU. This plot depicts the likelihood of discharge alive for varying ICU lengths of stay. The plot shows impact of having a clinical pharmacist providing CMM with rounding every day of ICU stay compared to patients who did not have CMM with rounding by a pharmacist at least 1 day of their ICU stay. CMM: comprehensive medication management, ICU: intensive care unit

All participating pharmacists held a Doctor of Pharmacy degree. Nearly all completed postgraduate training (97.1%) and held active board certification (94.2%). The mean years of practice post training was 7.5 ± 5.4 years. Regarding primary practice location, 11.6% of respondents practiced in the PICU, and 5.8% in the NICU, while most practiced in adult ICUs or as float pharmacists. Details on pharmacist characteristics are presented in **Table 1**. Pharmacist characteristics split by subgroup are provided in the **Supplementary Appendix**.

There were 786 patients admitted to PICUs, 563 to NICUs, and 110 to adult ICUs, the majority of whom were treated in an adult trauma ICU. The median patient age was 0.3 years (IQR 0-7.6), and 54.6% were male. The median PRISM III score was 3.0 (IQR 0-8.0), and the median MRC-ICU score was 5.0 (IQR 2.0-10.0). The median patient-specific average PPR was 25.8 (IQR 17.0-36.1). About a quarter of patients (23.0%) received daily CMM with rounding, while the remaining patients had at least one ICU day without pharmacist CMM with rounding, such as CMM without rounding. During hospitalization, 34.3% required mechanical ventilation, 15.9% required mechanical circulatory support, and 1.0% received renal replacement therapy. The overall mortality rate was 3.4%. The median ICU LOS was 4.0 days (IQR 2.0-13.0). The median hospital LOS was 6.4 days (3.0-17.0) and the median VFDs were 28.0 days (27-28). Additional patient characteristics are presented in **Table 2**.

**Table 2.** Patient characteristics and outcomes.

| Variable | Overall<br>(N=1,459) | No CMM with<br>rounding every<br>day (N=1,123) | CMM with<br>rounding every<br>day (N=336) | p-value |
| --- | --- | --- | --- | --- |
| Secondary Variable |  |  |  |  |
| Pharmacist-to-ICU-patient ratio | 25.8 (17.0, 36.1) | 28.0 (19.5, 39.0) | 16.0 (12.0, 26.0) | <0.001 |
| Missing | 20 (1.4) | 20 (1.8) | 0 (0) |  |
| Co-variables |  |  |  |  |
| Age, years | 0.3 (0.0, 7.6) | 0.1 (0.0, 5.7) | 2.9 (0.3, 11.3) | <0.001 |
| Sex |  |  |  | >0.9 |
| Male | 796 (54.6) | 611 (54.4) | 185 (55.1) |  |
| Female | 661 (45.3) | 510 (45.4) | 151 (44.9) |  |
| Unidentified | 2 (0.1) | 2 (0.2) | 0 (0.0) |  |
| MRC-ICU Score<sup>a</sup> | 5.0 (2.0, 10.0) | 5.0 (2.0, 10.0) | 3.0 (1.0, 8.0) | <0.001 |
| PRISM III Score<sup>a</sup> | 3.0 (0.0, 8.0) | 3.0 (0.0, 8.0) | 3.0 (0.0, 7.0) | 0.011 |
| Missing | 10 (0.7) | 4 (0.4) | 6 (1.8) |  |
| ICU admission day of the week |  |  |  | <0.001 |
| Monday | 198 (13.6) | 134 (11.9) | 64 (19.0) |  |
| Tuesday | 259 (17.8) | 167 (14.9) | 92 (27.4) |  |
| Wednesday | 238 (16.3) | 174 (15.5) | 64 (19.0) |  |
| Thursday | 222 (15.2) | 179 (15.9) | 43 (12.8) |  |
| Friday | 224 (15.4) | 208 (18.5) | 16 (4.8) |  |
| Saturday | 135 (9.3) | 127 (11.3) | 8 (2.4) |  |
| Sunday | 183 (12.5) | 134 (11.9) | 49 (14.6) |  |
| Institution type |  |  |  | <0.001 |
| Academic Medical Center | 1,222 (83.8) | 916 (81.6) | 306 (91.1) |  |
| Community Hospital | 237 (16.2) | 207 (18.4) | 30 (8.9) |  |
| ICU type |  |  |  | <0.001 |
| Pediatric | 786 (53.9) | 540 (48.1) | 246 (73.2) |  |
| Neonatal | 563 (38.6) | 513 (45.7) | 50 (14.9) |  |
| Adult Trauma | 64 (4.4) | 43 (3.8) | 21 (6.3) |  |
| Adult Neurosurgery/Neurology | 15 (1.0) | 14 (1.2) | 1 (0.3) |  |
| Adult Mixed Medical-Surgical | 17 (1.2) | 5 (0.4) | 12 (3.6) |  |
| Other Adult ICU | 14 (1.0) | 8 (0.7) | 6 (1.8) |  |
| ICU pharmacist coverage first 24 hours<sup>b</sup> |  |  |  | <0.001 |
| CMM delivered on interprofessional rounds | 966 (66.2) | 642 (57.2) | 336 (100) |  |
| CMM delivered not on interprofessional rounds | 192 (13.2) | 186 (16.6) | 0 (0.0) |  |
| Abbreviated CMM delivered | 67 (4.6) | 65 (5.8) | 0 (0.0) |  |
| CMM not delivered (absence of CMM) | 229 (15.7) | 225 (20.0) | 0 (0.0) |  |
| Unknown | 5 (0.3) | 5 (0.4) | 0 (0.0) |  |
| **Nurse-to-patient ratio (average), 1:X** |  |  |  | <0.001 |
| ≤1 | 107 (7.5) | 77 (7.0) | 30 (9.1) |  |
| 2 | 771 (53.9) | 531 (48.3) | 240 (72.5) |  |
| >2 | 252 (17.6) | 222 (20.2) | 30 (9.1) |  |
| 1-2 | 301 (21.0) | 270 (24.5) | 31 (9.4) |  |
| Missing | 28 (1.9) | 23 (2.0) | 5 (1.5) |  |
| **Percentage of medical teams rounded with<sup>c</sup>** | 0.6 (0.5, 1.0) | 0.6 (0.5, 0.8) | 0.8 (0.5, 1.0) | <0.001 |
| Missing | 25 (1.7) | 20 (1.8) | 5 (1.5) |  |
| **Renal replacement therapy** |  |  |  | >0.9 |
| No | 1,444 (99.0) | 1,111 (98.9) | 333 (99.1) |  |
| Yes | 15 (1.0) | 12 (1.1) | 3 (0.9) |  |
| **Mechanical circulatory support** |  |  |  | 0.022 |
| No | 1,227 (84.1) | 931 (82.9) | 296 (88.1) |  |
| Yes | 232 (15.9) | 192 (17.1) | 40 (11.9) |  |
| **Mechanical ventilation** |  |  |  | <0.001 |
| No | 959 (65.7) | 692 (61.6) | 267 (79.5) |  |
| Yes | 500 (34.3) | 431 (38.4) | 69 (20.5) |  |
| **Outcomes** |  |  |  |  |
| **Mortality** | 50 (3.4) | 43 (3.8) | 7 (2.1) | 0.12 |
| **ICU length of stay, days** | 4.0 (2.0, 13.0) | 6.8 (2.8, 18.0) | 1.7 (1.0, 2.5) | <0.001 |
| **ICU-free days** | 23.9 (14.0, 26.0) | 21.0 (9.0, 25.2) | 26.2 (25.1, 27.0) | <0.001 |
| Missing | 19 (1.3) | 18 (1.6) | 1 (0.3) |  |
| **Duration of mechanical ventilation, days** | 0.0 (0.0, 0.7) | 0.0 (0.0, 1.1) | 0.0 (0.0, 0.0) | <0.001 |
| **Ventilator-free days** | 28.0 (27.2, 28.0) | 28.0 (26.8, 28.0) | 28.0 (28.0, 28.0) | <0.001 |
| **Hospital length of stay, days** | 6.4 (3.0, 17.0) | 8.9 (3.9, 21.7) | 3.0 (1.9, 4.7) | <0.001 |

Data reported as n (%) or median (interquartile range)
CMM: comprehensive Medication Management; ICU: Intensive Care Unit; MRC-ICU: medication regimen complexity-intensive care unit; PRISM III: Pediatric Risk of Mortality III
<sup>a</sup>worst score during the first 24 hours of ICU stay
<sup>b</sup>CMM delivered on interprofessional rounds: the pharmacist attended multidisciplinary rounds and verbally provided CMM (including review of medications and recommendations); CMM delivered not on interprofessional rounds: the pharmacist provided CMM (reviewed medications and provided recommendations) but did not attend interdisciplinary rounds; Abbreviated CMM delivered: the pharmacist provided abbreviated CMM which includes brief medication review but does not include full patient review (e.g. progress notes, results) and recommendations were provided outside of rounds; CMM not delivered (absence of CMM): the patient received no medication review from a pharmacist outside of pharmacokinetic monitoring and prospective medication order verification
<sup>c</sup>Percent of assigned ICU team's pharmacist rounded is calculated by total number of ICU medical teams the pharmacist attended and provided CMM on interprofessional rounds for divided by total number of ICU medical teams providing care for patients that the pharmacist was assigned to provide CMM for

### Primary outcome (**Table 3**)

**Table 3.**
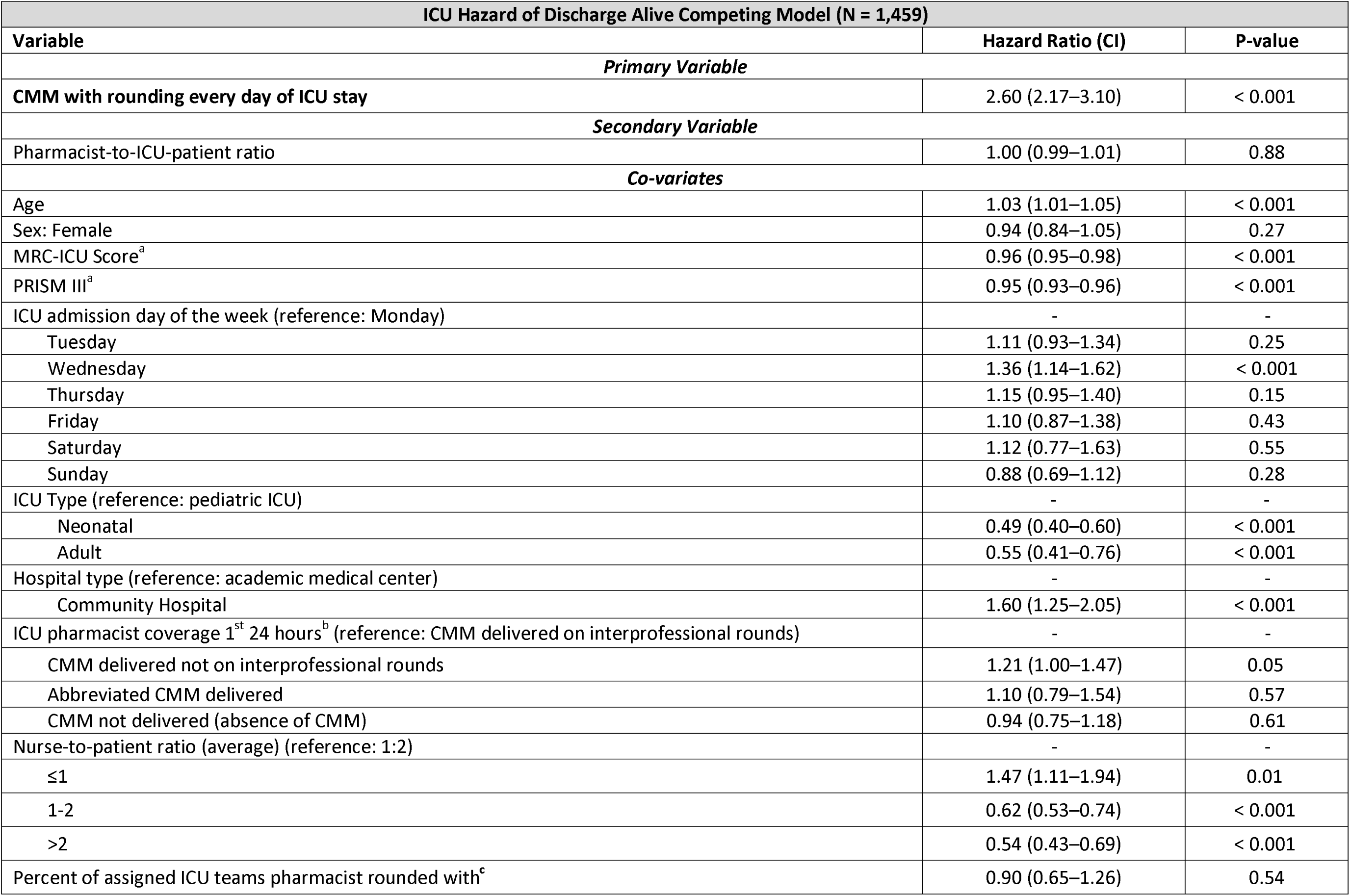
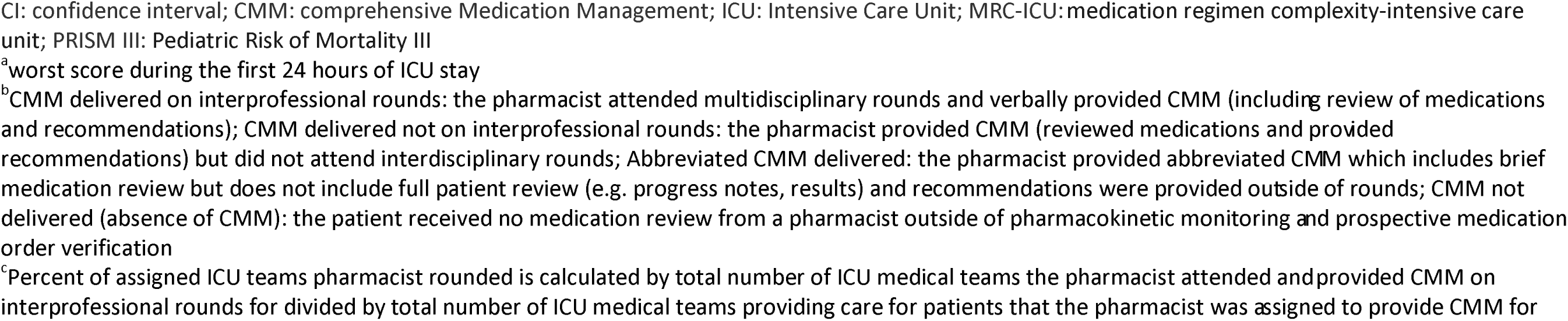
Fine-Gray Competing Risks Model for ICU Hazard of Discharge Alive with In-ICU Mortality as a Competing Event Based on Imputed Data.

| ICU Hazard of Discharge Alive Competing Model (N = 1,459) |  |  |
| --- | --- | --- |
| Variable | Hazard Ratio (CI) | P-value |
| *Primary Variable* |  |  |
| **CMM with rounding every day of ICU stay** | 2.60 (2.17–3.10) | < 0.001 |
| *Secondary Variable* |  |  |
| Pharmacist-to-ICU-patient ratio | 1.00 (0.99–1.01) | 0.88 |
| *Co-variables* |  |  |
| Age | 1.03 (1.01–1.05) | < 0.001 |
| Sex: Female | 0.94 (0.84–1.05) | 0.27 |
| MRC-ICU Score<sup>a</sup> | 0.96 (0.95–0.98) | < 0.001 |
| PRISM III<sup>a</sup> | 0.95 (0.93–0.96) | < 0.001 |
| ICU admission day of the week (reference: Monday) | - | - |
| Tuesday | 1.11 (0.93–1.34) | 0.25 |
| Wednesday | 1.36 (1.14–1.62) | < 0.001 |
| Thursday | 1.15 (0.95–1.40) | 0.15 |
| Friday | 1.10 (0.87–1.38) | 0.43 |
| Saturday | 1.12 (0.77–1.63) | 0.55 |
| Sunday | 0.88 (0.69–1.12) | 0.28 |
| ICU Type (reference: pediatric ICU) | - | - |
| Neonatal | 0.49 (0.40–0.60) | < 0.001 |
| Adult | 0.55 (0.41–0.76) | < 0.001 |
| Hospital type (reference: academic medical center) | - | - |
| Community Hospital | 1.60 (1.25–2.05) | < 0.001 |
| ICU pharmacist coverage 1<sup>st</sup> 24 hours<sup>b</sup> (reference: CMM delivered on interprofessional rounds) | - | - |
| CMM delivered not on interprofessional rounds | 1.21 (1.00–1.47) | 0.05 |
| Abbreviated CMM delivered | 1.10 (0.79–1.54) | 0.57 |
| CMM not delivered (absence of CMM) | 0.94 (0.75–1.18) | 0.61 |
| Nurse-to-patient ratio (average) (reference: 1:2) | - | - |
| ≤1 | 1.47 (1.11–1.94) | 0.01 |
| 1-2 | 0.62 (0.53–0.74) | < 0.001 |
| >2 | 0.54 (0.43–0.69) | < 0.001 |
| Percent of assigned ICU teams pharmacist rounded with<sup>c</sup> | 0.90 (0.65–1.26) | 0.54 |

CI: confidence interval; CMM: comprehensive Medication Management; ICU: Intensive Care Unit; MRC-ICU: medication regimen complexity-intensive care unit; PRISM III: Pediatric Risk of Mortality III
<sup>a</sup>worst score during the first 24 hours of ICU stay
<sup>b</sup>CMM delivered on interprofessional rounds: the pharmacist attended multidisciplinary rounds and verbally provided CMM (including review of medications and recommendations); CMM delivered not on interprofessional rounds: the pharmacist provided CMM (reviewed medications and provided recommendations) but did not attend interdisciplinary rounds; Abbreviated CMM delivered: the pharmacist provided abbreviated CMM which includes brief medication review but does not include full patient review (e.g. progress notes, results) and recommendations were provided outside of rounds; CMM not delivered (absence of CMM): the patient received no medication review from a pharmacist outside of pharmacokinetic monitoring and prospective medication order verification
<sup>c</sup>Percent of assigned ICU teams pharmacist rounded is calculated by total number of ICU medical teams the pharmacist attended and provided CMM on interprofessional rounds for divided by total number of ICU medical teams providing care for patients that the pharmacist was assigned to provide CMM for

In the Fine-Gray competing risk model for the primary outcome of ICU HAD, patients receiving daily CMM with rounding had 160% higher HDA from the ICU compared with those who did not (HR 2.60, 95% CI 2.17-3.10; p<0.001). The PPR was not significantly associated with the ICU HDA (HR 1.00, 95% CI 0.99-1.01; p = 0.88).

### Secondary Outcomes (**Table 4**)

In adjusted Firth logistic regression, daily CMM with rounding was not significantly associated with overall in-hospital mortality (OR 0.44, 95% CI 0.14-1.36; p=0.15). Patients who received daily CMM with rounding had a 125% higher HDA from the hospital (HR 2.25, 95% CI 1.83-2.75; p<0.001) and a 21% higher HEA (HR 1.21, 95% CI 1.10-1.33; p<0.001). When the PPR was evaluated as the secondary exposure, no significant associations were observed with any secondary outcome, including in-hospital mortality (OR 1.03, 95% CI 0.98–1.08; p = 0.304), hospital HDA (HR 1.01, 95% CI 1.00–1.02; p = 0.309), or HEA (HR 1.00, 95% CI 0.99–1.00; p = 0.223).

**Table 4:** Regression Analyses for Mortality and Fine-Gray Competing Risks Model for Hospital Hazard of Discharge Alive and HEA with in-ICU Mortality as a Competing Event.

| Variable | In-hospital Mortality |  |  |  | Hazard of Discharge Alive from Hospital |  | Hazard of Extubation Alive |  |
| --- | --- | --- | --- | --- | --- | --- | --- | --- |
|  | Univariable<br>OR (CI) | P-value | Multivariable<br>OR (CI) | P-value | HR (CI) | P-value | HR (CI) | P-value |
| *Primary Variable* |  |  |  |  |  |  |  |  |
| **CMM with rounding every day of ICU stay** | 0.57 (0.26, 1.24) | 0.15 | 0.44 (0.14, 1.36) | 0.15 | 2.25 (1.83–2.75) | < 0.001 | 1.21 (1.10–1.34) | < 0.001 |
| *Secondary Variable* |  |  |  |  |  |  |  |  |
| **Pharmacist-to-ICU-patient ratio** | 0.99 (0.96, 1.01) | 0.24 | 1.03 (0.98, 1.08) | 0.30 | 1.01 (1.00–1.02) | 0.31 | 1.00 (0.99–1.00) | 0.22 |
| *Co-variables* |  |  |  |  |  |  |  |  |
| **Age** | 1.03 (0.99, 1.08) | 0.12 | 0.92 (0.84, 1.01) | 0.07 | 1.02 (1.00–1.04) | 0.02 | 1.02 (1.01–1.03) | < 0.001 |
| **Sex: female** | 0.87 (0.50, 1.54) | 0.64 | 0.94 (0.50, 1.78) | 0.85 | 0.97 (0.86–1.09) | 0.61 | 1.02 (0.95–1.10) | 0.56 |
| **MRC-ICU Score<sup>a</sup>** | 1.15 (1.11, 1.19) | <0.001 | 1.05 (1.01, 1.11) | 0.03 | 0.96 (0.95–0.97) | < 0.001 | 0.95 (0.94–0.96) | < 0.001 |
| **PRISM III<sup>a</sup>** | 1.23 (1.18, 1.28) | <0.001 | 1.16 (1.11, 1.21) | <0.001 | 0.93 (0.92–0.95) | < 0.001 | 0.95 (0.94–0.96) | < 0.001 |
| **ICU admission day of the week (reference: Monday)** | - | - | - | - | - | - | - | - |
| Tuesday | 0.38 (0.15, 1.01) | 0.05 | 0.40 (0.13, 1.25) | 0.12 | 1.17 (0.94–1.46) | 0.15 | 1.14 (0.99–1.31) | 0.08 |
| Wednesday | 0.35 (0.13, 0.98) | 0.05 | 0.47 (0.15, 1.49) | 0.20 | 1.27 (1.02–1.58) | 0.03 | 1.15 (1.00–1.32) | 0.06 |
| Thursday | 0.24 (0.07, 0.79) | 0.02 | 0.34 (0.10, 1.13) | 0.08 | 1.16 (0.93–1.44) | 0.19 | 1.14 (0.98–1.33) | 0.09 |
| Friday | 0.80 (0.35, 1.83) | 0.60 | 0.92 (0.31, 2.72) | 0.88 | 1.00 (0.77–1.30) | 1.00 | 1.10 (0.91–1.32) | 0.34 |
| Saturday | 0.75 (0.28, 1.98) | 0.56 | 0.73 (0.19, 2.81) | 0.65 | 1.04 (0.74–1.46) | 0.83 | 1.13 (0.89–1.44) | 0.30 |
| Sunday | 0.63 (0.25, 1.60) | 0.34 | 1.15 (0.37, 3.54) | 0.81 | 0.90 (0.70–1.16) | 0.41 | 0.99 (0.84–1.18) | 0.94 |
| **ICU Type (reference: pediatric ICU)** | - | - | - | - | - | - | - | - |
| Neonatal | 1.07 (0.56, 2.06) | 0.83 | 0.69 (0.24, 2.02) | 0.50 | 0.52 (0.42–0.63) | < 0.001 | 1.05 (0.92–1.20) | 0.50 |
| Adult | 4.93 (2.42, 10.05) | <0.001 | 6.42 (1.40, 29.51) | 0.02 | 0.45 (0.32–0.63) | < 0.001 | 0.61 (0.50–0.75) | < 0.001 |
| **Hospital type (reference: academic medical center)** | - | - | - | - | - | - | - | - |
| Community Hospital | 1.71 (0.89, 3.28) | 0.11 | 4.20 (1.39, 12.68 ) | 0.01 | 1.33 (1.00–1.77) | 0.05 | 1.05 (0.85–1.29) | 0.65 |
| **ICU pharmacist coverage 1<sup>st</sup> 24 hours<sup>b</sup>**<br>**(reference: CMM delivered on**<br>**interprofessional rounds)** | - | - | - | - | - | - | - | - |
| CMM delivered not on interprofessional rounds | 1.00 (0.42, 2.35) | 1.00 | 1.04 (0.36, 3.04) | 0.94 | 1.21 (1.00–1.46) | 0.05 | 1.04 (0.92–1.17) | 0.57 |
| Abbreviated CMM delivered | 1.10 (0.30, 4.06) | 0.89 | 0.81 (0.17, 3.78) | 0.79 | 0.85 (0.62–1.17) | 0.31 | 0.93 (0.73–1.18) | 0.54 |
| CMM not delivered (absence of CMM) | 1.38 (0.68, 2.81) | 0.38 | 1.30 (0.47, 3.59) | 0.62 | 1.04 (0.84–1.29) | 0.70 | 0.93 (0.79–1.10) | 0.40 |
| **Nurse-to-patient ratio (average)**<br>**(reference: 1:2)** | - | - | - | - | - | - | - | - |
| ≤1 | 5.31 (2.36, 11.95) | <0.001 | 5.04 (1.66, 15.31 ) | 0.004 | 0.85 (0.63–1.17) | 0.32 | 0.95 (0.82–1.09) | 0.45 |
| 1-2 | 4.00 (2.07, 7.75) | <0.001 | 2.71 (1.22, 6.01) | 0.01 | 0.54 (0.45–0.63) | < 0.001 | 0.76 (0.67–0.86) | < 0.001 |
| >2 | 0.70 (0.22, 2.24) | 0.54 | 0.15 (0.03, 0.66) | 0.01 | 0.62 (0.48–0.80) | < 0.001 | 1.11 (0.92–1.34) | 0.28 |
| **Percent of assigned ICU teams**<br>**pharmacist rounded with<sup>c</sup>** | 0.68 (0.22, 2.09) | 0.50 | 1.22 (0.21, 7.13) | 0.83 | 0.76 (0.50–1.14) | 0.18 | 0.89 (0.73–1.10) | 0.29 |

CI: confidence interval; CMM: comprehensive Medication Management; HR: hazard ratio; ICU: Intensive Care Unit; MRC-ICU: medication regimen complexity-intensive care unit; OR: odds ratio; PRISM III: Pediatric Risk of Mortality III
<sup>a</sup>worst score during the first 24 hours of ICU stay
<sup>b</sup>CMM delivered on interprofessional rounds: the pharmacist attended multidisciplinary rounds and verbally provided CMM (including review of medications and recommendations); CMM delivered not on interprofessional rounds: the pharmacist provided CMM (reviewed medications and provided recommendations) but did not attend interdisciplinary rounds; Abbreviated CMM delivered: the pharmacist provided abbreviated CMM which includes brief medication review but does not include full patient review (e.g. progress notes, results) and recommendations were provided outside of rounds; CMM not delivered (absence of CMM): the patient received no medication review from a pharmacist outside of pharmacokinetic monitoring and prospective medication order verification
<sup>c</sup>Percent of assigned ICU teams pharmacist rounded is calculated by total number of ICU medical teams the pharmacist attended and provided CMM on interprofessional rounds for divided by total number of ICU medical teams providing care for patients that the pharmacist was assigned to provide CMM for

### PICU Subgroup

Details on subgroup analyses are in the **Supplementary Appendix**. Among the 786 PICU patients, the median age was 2.9 years (IQR 0.5–10.2), 53.3% were male, and 21 patients (2.7%) died during hospitalization. Daily CMM with rounding was reported for 31.3% of patients and the median PPR was 19.0 (IQR 14.0-26.4). The median LOS in the ICU and hospital was 2.7 days (IQR 1.0–5.3) and 4.2 days (IQR 2.5–8.8), respectively, and VFDs were 28.0 days (IQR 27.3–28.0). In adjusted analyses, patients receiving daily CMM with rounding demonstrated a significantly higher ICU HDA (HR 2.29, 95% CI 1.91–2.74; p<0.001) and hospital HDA (HR 1.99, 95% CI 1.61–2.46, p<0.001). Daily CMM with rounding was also associated with increased HEA (HR 1.38, 95% CI 1.23–1.54, p<0.001) and lower odds of in-hospital mortality (OR 0.15, 95% CI 0.03–0.79, p=0.025). A higher PPR ratio was associated with a slightly lower ICU HDA (HR 0.99, 95% CI 0.98–1.00; p=0.03) and a slightly lower HEA (HR 0.99, 95% CI, 0.99–1.00; p<0.001) but was not significantly associated with the hospital HDA (HR 1.00, 95% CI 1.00–1.01; p=0.80) or in-hospital mortality (OR 1.06, 95% CI 1.00–1.14; p=0.11).

### NICU Subgroup

Among the 563 NICU patients, 53.6% were male, and 16 patients (2.8%) died during hospitalization. Daily CMM with rounding was less common than in the PICU cohort, occurring in 8.9% of patients, and the median PPR was 38.2 (IQR 30.5-44.0). Median ICU LOS was 12.0 days (IQR 5.0–35.0), median hospital LOS was 12.6 days (IQR 5.5–37.2), and median VFDs were 28.0 days (IQR 27.9–28.0). In adjusted analyses, daily CMM with rounding was associated with a significantly higher likelihood of ICU discharge (HR 5.78, 95% CI 3.92–8.53, p<0.001) and hospital discharge (HR 5.67, 95% CI 3.41–9.42; p<0.001). However, no significant associations were observed between daily CMM with rounding and mortality (OR 1.08, 95% CI 0.09–12.99; p=0.95) or VFDs (HR 0.98, 95% CI 0.84–1.14; p=0.77). A higher PPR was associated with a higher ICU HDA (HR 1.03, 95% CI 1.01–1.04; p=0.008) but was not significantly associated with the hospital HDA (HR 1.02, 95% CI 1.00–1.04; p=0.09), the HEA (HR 1.00, 95% CI, 0.99–1.01; p 0.46), or in-hospital mortality (OR 1.01, 95% CI 0.93–1.10; p=0.75).

## DISCUSSION

In this multicenter observational study, pharmacist-provided CMM with daily ICU rounding was associated with improved patient outcomes, including significantly higher ICU and hospital HDAs as well as increased VFDs. In subgroup analyses, CMM with daily ICU rounding was linked to higher ICU and hospital HDAs in both PICU and NICU populations. Although the association with mortality did not reach statistical significance in the overall cohort, the PICU subgroup demonstrated a significant reduction in in-hospital mortality among patients receiving daily CMM with rounding. The insignificant findings for mortality and VFDs in neonates may be attributable to the relatively small number of events and lower prevalence of daily CMM exposure in this population.

These results are consistent with adult ICU literature showing that pharmacist involvement in daily multidisciplinary rounding and CMM improves patient outcomes.^13–15,17–22,28–37^ The consistency across age groups suggests that the mechanisms by which pharmacists improve outcomes are fundamental to critical care practice: real-time medication optimization, early ADE identification, and prompt therapy adjustment in response to clinical changes. Similar trends have been observed in previous pediatric studies showing that pharmacist clinical involvement increases interventions and reduces ADEs, further supporting the value of integrating pharmacists into daily multidisciplinary ICU patient care.^2,3^

The PPR was not associated with clinical outcomes in the overall cohort, suggesting that pharmacist staffing alone may not adequately capture the impact of clinical pharmacist involvement. Although studies in adult ICU populations have reported associations between pharmacist staffing ratios and mortality,^23^ differences in practice models and workload allocation may explain these differences. Further research should evaluate staffing within defined clinical services to better understand how staffing models impact patient outcomes. It is noted that while subgroup analyses demonstrated statistically significant associations between PPR and ICU HDA in PICU and NICU patients, the effect sizes were small, and the direction of association differed between the two populations. The substantial difference in median PPR between PICU and NICU settings suggests distinct patient acuity and pharmacist practice models. Given the absence of significant associations overall and inconsistent subgroup findings, these results should be interpreted cautiously and may represent limited power rather than clinically meaningful differences. Another notable finding was that 79.7% of pharmacists care for only 7.5% of pediatric patients in adult ICUs, reflecting shared or float-based coverage rather than dedicated pediatric staffing. This subgroup was not analyzed separately and may differ from patients cared for in PICUs and NICUs, warranting further investigation.

At present, clinical pharmacy practice in the NICU and PICU varies across institutions and is often constrained by competing demands between direct and indirect patient care responsibilities.^5,14,15^ Findings from this study support standardized pediatric pharmacy practice models that define expectations for the pharmacist to participate in rounds on a daily basis, including weekends, and provide CMM. Incorporating these evidence-based pharmacy clinical services into standard workflow may enhance medication safety, optimize medication therapy, and improve outcomes for critically ill children.

This is the first multicenter observational study showing the impact of pharmacy services and patient outcomes in the non-adult ICU setting. While previous studies show positive impacts of CMM services on patient outcomes in adults,^16–22^ this study extend the existing literature by demonstrating associations between CMM daily with multidisciplinary round participation and selected clinical outcomes in PICU and NICU settings. The multicenter design enhances the generalizability of the study findings. Several limitations should be acknowledged. As an observational study, residual confounding factors cannot be fully excluded despite adjustment for multiple confounding variables. For example, outcomes may be affected by weekend and off-hour pharmacist interventions that were not captured in this study. Institutional characteristics, such as academic center and the capacity to maintain lower staffing ratios, may contribute to the observed outcomes. Additionally, the relatively small number of mortality events, particularly in the NICU subgroup, limited the ability to detect differences in rare outcomes. Differences in patient characteristics between PICU and NICU populations may contribute to variation in outcomes, with PICU patients potentially representing a more heterogenous population with greater variation in medical complexity; however, these population-level differences could not be adequately examined in this study. Further investigation of pharmacist interventions within each population with larger cohorts and standardized pharmacist interventions may be warranted to better understand clinical pharmacist intervention impact on PICU and NICU patients.

## CONCLUSIONS

Pharmacist-provided daily CMM with ICU rounding is associated with higher ICU and hospital HDA in both PICU and NICU populations. These findings provide evidence supporting the integration of clinical pharmacists into daily multiprofessional rounds and the delivery of CMM as a standardized pharmacy practice in the PICU and NICU. Results from this study inform staffing models and operational priorities, emphasizing the need for institutions to allocate pharmacist resources in ways that ensure consistent delivery of these services to improve patient outcomes.

## Data Availability

All data produced in the present study are available upon reasonable request to the authors

## Contributor Roles Taxonomy (CRediT)

All authors contributed to the research and manuscript and meet ICJME authorship criteria. SES had full access to all of the data in the study and takes responsibility for the integrity of the data and the accuracy of the data analysis, including and especially any adverse effects. SES, KRH, MH, KB, and AS contributed substantially to the study design, data analysis and interpretation, and the writing of the manuscript. ZC and XC conducted statistical analysis and interpretation and participated in writing the first draft of the manuscript. AK, JM, HTD, and EF provided study design oversight and provided feedback on data interpretation and the manuscript. The OPTIM Investigator Team completed data collection and provided feedback on data interpretation and the manuscript.

## PRESUBMISSION CONSULTATION

: proposed submission and explanation of its significance

Clinical pharmacists play an important role in optimizing medication therapy and promoting medication safety in critically ill patients, yet standardized models for pharmacist integration in pediatric intensive care remain limited. This multicenter study provides evidence that daily pharmacist-led comprehensive medication management with participation in multidisciplinary ICU rounds is associated with improved patient outcomes, including higher odds of ICU and hospital discharge alive and extubation alive. These findings support consistent integration of clinical pharmacists into daily multidisciplinary rounds in PICU and NICU settings and may inform the development of standardized pediatric clinical pharmacy practice models and staffing strategies.

summary (for social media communication, promotion)

Standardized pediatric critical care pharmacy models are lacking. This multicenter study found that daily pharmacist-led medication management and rounding were associated with improved outcomes, supporting consistent pharmacist integration into multiprofessional care teams.

Report in Context

- Increased clinical pharmacist involvement in NICUs and PICUs is associated with improved patient care. However, the specific clinical pharmacy services that contribute to these positive outcomes remain poorly understood.
- Comprehensive medication management (CMM) has become the standard of care in the adult ICU given its proven benefits in preventing medication adverse events and reducing mortality. However, the impact of CMM in pediatric populations is not well established.
- While a growing body of literature underscores that a multiprofessional ICU approach including clinical pharmacists improves patient outcomes, few studies examine clinical pharmacists’ direct contribution to patient outcomes, particularly in neonatal and pediatric critical care.

At the Bedside

- Pharmacist-provided CMM with daily ICU rounding was associated with improved outcomes, including higher ICU and hospital HDAs and more VFDs, consistent with adult ICU evidence and supporting the importance of clinical pharmacist services across critically ill populations.
- Differing from adult ICUs, PPR was not associated with outcomes except in PICUs. Differences in practice models, workload allocation, direct patient care time, and team integration may contribute; further research should evaluate staffing within defined service models.
- Standardized clinical pharmacy practice models are lacking in NICUs and PICUs. These findings support models defining expectations for daily pharmacist participation in multidisciplinary rounds and provision of CMM to optimize patient care.

## REFERENCES

1. Sikora A, Murray B, Henry K, et al. Consensus recommendations for the integration of critical care pharmacists on intensive care unit teams: Endorsed by the American Association of Critical-Care Nurses, American College of Clinical Pharmacy, American Society of Health-System Pharmacists, Institute for Safe Medication Practices, and Society of Critical Care Medicine. JACCP. 2025;8(9) 914–930. doi:doi:10.1002/jac5.70084

2. Tripathi S, Crabtree HM, Fryer KR, Graner KK, Arteaga GM. Impact of Clinical Pharmacist on the Pediatric Intensive Care Practice: An 11-Year Tertiary Center Experience. J Pediatr Pharmacol Ther. Jul–Aug 2015;20(4):290–8. doi:10.5863/1551-6776-20.4.290

3. Blowey B, Resendiz KV, Grachen A, Srinivasan V. Prevention is better than cure: The vital role of the clinical pharmacist in the pediatric intensive care unit to prevent medication errors. Journal of Pediatric Critical Care. 2020;7(5):235–236. doi:10.4103/jpcc.Jpcc_110_20

4. Holden DN, Groth CM, Acquisto NM, et al. Measuring Critical Care Pharmacist Value: A Scoping Review of the Effect of Critical Care Pharmacist Activities on Clinically Relevant Outcomes. JACCP. 2026;3:370188.

5. Buckley MS, Smith SE, Birriel B, et al. ICU Patient-to-Pharmacist Ratios: A Prospective, Multicenter Time-Motion Study. Crit Care Med. Apr 1 2025;53(4):e863–e873. doi:10.1097/ccm.0000000000006605

6. Yang CP, Veltri MA, Anton B, Yaster M, Berkowitz ID. Food and Drug Administration approval for medications used in the pediatric intensive care unit: a continuing conundrum. Pediatr Crit Care Med. Sep 2011;12(5):e195–9. doi:10.1097/PCC.0b013e3181fe25b9

7. Czaja AS, Reiter PD, Schultz ML, Valuck RJ. Patterns of Off-Label Prescribing in the Pediatric Intensive Care Unit and Prioritizing Future Research. J Pediatr Pharmacol Ther. May–Jun 2015;20(3):186–96. doi:10.5863/1551-6776-20.3.186

8. Kaushal R, Bates DW, Landrigan C, et al. Medication Errors and Adverse Drug Events in Pediatric Inpatients. JAMA. 2001;285(16):2114–2120. doi:10.1001/jama.285.16.2114

9. Eiland LS, Benner K, Gumpper KF, et al. ASHP-PPAG Guidelines for Providing Pediatric Pharmacy Services in Hospitals and Health Systems. J Pediatr Pharmacol Ther. May–Jun 2018;23(3):177–191. doi:10.5863/1551-6776-23.3.177

10. Comprehensive Medication Management: promoting optimized and personalized use of medication through comprehensive medication management. Accessed 06/15, 2026. https://www.ashp.org/advocacy-and-issues/key-issues/other-issues/comprehensive-medication-management?loginreturnUrl=SSOCheckOnly

11. American Society of Health-System Pharmacists. Comprehensive Medication Management. Accessed May 12, 2026. https://www.ashp.org/advocacy-and-issues/key-issues/other-issues/comprehensive-medication-management?loginreturnUrl=SSOCheckOnly

12. Health OMfB. Comprehensive Medication Management. 2025. https://www.optimizingmeds.org/comprehensive-medication-management/

13. Sikora A, Min W, Devlin JW, et al. Effect of Comprehensive Medication Management on Mortality in Critically Ill Patients. Critical Care Medicine. 2025;53(10):e1995–e2004. doi:10.1097/ccm.0000000000006802

14. Smith SE, Henry KR, Heavner MS, et al. Optimization of Pharmacist Medication Management and Mortality in the Intensive Care Unit. CHEST. 2026;0 doi:10.1016/j.chest.2026.09.027

15. Henry KR, Smith BA, Holden DN, et al. Impact of Early Critical Care Pharmacist Involvement on Patient Outcomes in the Intensive Care Unit. medRxiv. 2026:2026.08.25.26361345. doi:10.64898/2026.08.25.26361345

16. Kim MM, Barnato AE, Angus DC, Fleisher LF, Kahn JM. The Effect of Multidisciplinary Care Teams on Intensive Care Unit Mortality. Archives of Internal Medicine. 2010;170(4):369–376. doi:10.1001/archinternmed.2009.521

17. Kim C, Durkin J, Dittmer A, et al. 1022: EVALUATING THE IMPACT OF EXPANDING DECENTRALIZED CLINICAL PHARMACY SERVICES TO THE MICU ON WEEKENDS. Critical Care Medicine. 2025/01/30 2025;53(1)doi:10.1097/01.ccm.0001102752.12443.c8

18. Kucukarslan SN, Corpus K, Mehta N, et al. Evaluation of a dedicated pharmacist staffing model in the medical intensive care unit. Hosp Pharm. Dec 2013;48(11):922–30. doi:10.1310/hpj4811-922

19. Kucukarslan SN, Peters M, Mlynarek M, Nafziger DA. Pharmacists on Rounding Teams Reduce Preventable Adverse Drug Events in Hospital General Medicine Units. Archives of Internal Medicine. 2003;163(17):2014–2018. doi:10.1001/archinte.163.17.2014

20. Leape LL, Cullen DJ, Clapp MD, et al. Pharmacist participation on physician rounds and adverse drug events in the intensive care unit. JAMA. Jul 21 1999;282(3):267–70. doi:10.1001/jama.282.3.267

21. Lee H, Ryu K, Sohn Y, Kim J, Suh GY, Kim E. Impact on Patient Outcomes of Pharmacist Participation in Multidisciplinary Critical Care Teams: A Systematic Review and Meta-Analysis. Crit Care Med. Sep 2019;47(9):1243–1250. doi:10.1097/CCM.0000000000003830

22. Montazeri M, Cook DJ. Impact of a clinical pharmacist in a multidisciplinary intensive care unit. Critical Care Medicine. 1994;22(6):1044–1048.

23. Smith SE, Henry KR, Heavner MS, et al. Optimization of Pharmacist Medication Management and Mortality in the Intensive Care Unit. medRxIV. 2026;doi:doi.org/10.64898/2025.12.31.25342855

24. Keats K, Sikora A, Heavner MS, Chen X, Smith SE, Team obotOI. Optimizing Pharmacist Team-Integration for ICU Patient Management: Rationale, Study Design, and Methods for a Multicentered Exploration of Pharmacist-to-Patient Ratio. Critical Care Explorations. 2023;5(9):e0956. doi:10.1097/cce.0000000000000956

25. REDCap-Reserach Electronic Data Capture. Accessed 06/17, 2026. https://projectredcap.org/about/

26. Pollack MM, Patel KM, Ruttimann UE. PRISM III: an updated Pediatric Risk of Mortality score. Crit Care Med. May 1996;24(5):743–52. doi:10.1097/00003246-199605000-00004

27. Gwynn ME, Poisson MO, Waller JL, Newsome AS. Development and validation of a medication regimen complexity scoring tool for critically ill patients. Am J Health Syst Pharm. May 17 2019;76(Supplement_2):S34–s40. doi:10.1093/ajhp/zxy054

28. Elrefaei H, El Nekidy WS, Nasef R, et al. The Impact of Clinical Pharmacist-Driven Weekend Antimicrobial Stewardship Coverage at a Quaternary Hospital. Antibiotics (Basel). Oct 16 2024;13(10)doi:10.3390/antibiotics13100974

29. Fraser T, Rivard K, Richter S, et al. The Implementation and Effect of Weekend Pharmacy-Driven Antimicrobial Stewardship Services at a Large Academic Medical Center. Journal of Pharmacy Practice. 2022/08/01 2022;35(4):541–545. doi:10.1177/0897190021997008

30. Kruer RM, Czosnowski Q, Miller EM, et al. Expansion of around the clock critical care and emergency medicine clinical pharmacy services at a large urban academic medical center. JACCP: JOURNAL OF THE AMERICAN COLLEGE OF CLINICAL PHARMACY. 2023;6(8):889–899. 10.1002/jac5.1850

31. Rech MA, Gurnani PK, Peppard WJ, et al. PHarmacist Avoidance or Reductions in Medical Costs in CRITically Ill Adults: PHARM-CRIT Study. Crit Care Explor. Dec 2021;3(12):e0594. doi:10.1097/CCE.0000000000000594

32. Sikora A, Ayyala D, Rech MA, et al. Impact of Pharmacists to Improve Patient Care in the Critically Ill: A Large Multicenter Analysis Using Meaningful Metrics With the Medication Regimen Complexity-ICU (MRC-ICU) Score. Crit Care Med. Sep 1 2022;50(9):1318–1328. doi:10.1097/CCM.0000000000005585

33. Sikora A MB, Most A, Martin G. Critical Care Pharmacists Save Lives. ICU Management and Practice. 2024;https://healthmanagement.org/c/icu/IssueArticle/critical-care-pharmacists-save-lives

34. Smetana KS, Flannery AH, Gurnani PK, et al. PHarmacist avoidance or reductions in medical costs in CRITically ill adults rounding with one SERVICE compared to two or more services: PHARM-CRIT-SERVICE. JACCP: JOURNAL OF THE AMERICAN COLLEGE OF CLINICAL PHARMACY. 2023;6(9):1000–1007. 10.1002/jac5.1798

35. Smith SE, Bektas, A. N., Henry, K., Heavner, M. S., Papdopoulos, J., Sikora, A. Optimizing team integration of critical care pharmacists: A history of critical care pharmacy practice and evolving definition of “optimal practice”. Critical Care Clinics. 2026;[in press]

36. Smith SE, Henry K, Heavner MS, et al. Impact of Pharmacist Board Certification on Health Outcomes of Critically Ill Patients: An Analysis of the Optimizing Pharmacist-Team Integration for ICU Patient Management (OPTIM) Study. JACCP: Journal of the American College of Clinical Pharmacy. 2026/07/07 2026;9(7)doi:10.1002/jac5.70258

37. Wang T, Benedict N, Olsen KM, et al. Effect of critical care pharmacist’s intervention on medication errors: a systematic review and meta-analysis of observational studies. Journal of critical care. 2015;30(5):1101–1106.

