## Supplemental Appendix for "Pharmacist-Led Comprehensive Medication Management and Patient Outcomes in Pediatric Intensive Care Unit Patients: A Multicenter Cohort Study"

### List of Optimizing Pharmacist Team-Integration for ICU Patient Management (OPTIM) Investigators

### List of Acknowledgements

### Reporting of Observational Studies in Epidemiology (STROBE) Checklist

|  | Item No | Recommendation | Page No |
| --- | --- | --- | --- |
| **Title and abstract** | 1 | (*a*) Indicate the study’s design with a commonly used term in the title or the abstract |  |
|  |  | (*b*) Provide in the abstract an informative and balanced summary of what was done and what was found |  |
| Introduction | | | |
| Background/rationale | 2 | Explain the scientific background and rationale for the investigation being reported |  |
| Objectives | 3 | State specific objectives, including any prespecified hypotheses |  |
| Methods | | | |
| Study design | 4 | Present key elements of study design early in the paper |  |
| Setting | 5 | Describe the setting, locations, and relevant dates, including periods of recruitment, exposure, follow-up, and data collection |  |
| Participants | 6 | (*a*) Give the eligibility criteria, and the sources and methods of selection of participants. Describe methods of follow-up |  |
|  |  | (*b*) For matched studies, give matching criteria and number of exposed and unexposed |  |
| Variables | 7 | Clearly define all outcomes, exposures, predictors, potential confounders, and effect modifiers. Give diagnostic criteria, if applicable |  |
| Data sources/ measurement | 8* | For each variable of interest, give sources of data and details of methods of assessment (measurement). Describe comparability of assessment methods if there is more than one group |  |
| Bias | 9 | Describe any efforts to address potential sources of bias |  |
| Study size | 10 | Explain how the study size was arrived at |  |
| Quantitative variables | 11 | Explain how quantitative variables were handled in the analyses. If applicable, describe which groupings were chosen and why |  |
| Statistical methods | 12 | (*a*) Describe all statistical methods, including those used to control for confounding |  |
|  |  | (*b*) Describe any methods used to examine subgroups and interactions |  |
|  |  | (*c*) Explain how missing data were addressed |  |
|  |  | (*d*) If applicable, explain how loss to follow-up was addressed |  |
|  |  | (*e*) Describe any sensitivity analyses |  |
| Results | | |  |
| Participants | 13* | (a) Report numbers of individuals at each stage of study—eg numbers potentially eligible, examined for eligibility, confirmed eligible, included in the study, completing follow-up, and analysed |  |
|  |  | (b) Give reasons for non-participation at each stage |  |
|  |  | (c) Consider use of a flow diagram |  |
| Descriptive data | 14* | (a) Give characteristics of study participants (eg demographic, clinical, social) and information on exposures and potential confounders |  |
|  |  | (b) Indicate number of participants with missing data for each variable of interest |  |
|  |  | (c) Summarise follow-up time (eg, average and total amount) |  |
| Outcome data | 15* | Report numbers of outcome events or summary measures over time |  |

| Main results | 16 | (*a*) Give unadjusted estimates and, if applicable, confounder-adjusted estimates and their precision (eg, 95% confidence interval). Make clear which confounders were adjusted for and why they were included |
| --- | --- | --- |
|  |  | (*b*) Report category boundaries when continuous variables were categorized |
|  |  | (*c*) If relevant, consider translating estimates of relative risk into absolute risk for a meaningful time period |
| Other analyses | 17 | Report other analyses done—eg analyses of subgroups and interactions, and sensitivity analyses |
| Discussion | | |
| Key results | 18 | Summarise key results with reference to study objectives |
| Limitations | 19 | Discuss limitations of the study, taking into account sources of potential bias or imprecision. Discuss both direction and magnitude of any potential bias |
| Interpretation | 20 | Give a cautious overall interpretation of results considering objectives, limitations, multiplicity of analyses, results from similar studies, and other relevant evidence |
| Generalisability | 21 | Discuss the generalisability (external validity) of the study results |
| Other information | | |
| Funding | 22 | Give the source of funding and the role of the funders for the present study and, if applicable, for the original study on which the present article is based |

*Give information separately for exposed and unexposed groups.

### Disclosures

### Additional Methods

#### eMethods 1. Definition of Pharmacist-to-Patient Ratio

The daily pharmacist-to-patient ratio was calculated for each ICU patient and averaged over the course of their ICU stay. For example, if a patient had a 3-day ICU length of stay and the pharmacist caring for the patient also covered 12, 8, and 14 ICU patients across these three days, then the patient-specific pharmacist-to-patient ratio would be: (12+8+14)/3 = 11.3 (ratio: 1:11). The daily pharmacist-patient ratio (all patients) was also calculated each day and included all patients (ICU as well as floor or step-down patients) that the pharmacist was assigned to provide CMM for. If the patient did not receive CMM, that day did not count towards their calculation of the pharmacist-to-patient ratio, as any attempt to quantify the effective ratio for no CMM would have been pure speculation. Thus, not receiving CMM was treated as a separate variable for analysis to distinguish these patients, as the care they received is likely not truly reflected by the ratio alone. For example, if a patient had a 4-day ICU length of stay and the pharmacist caring for the patient also covered 12, 8, and 14 ICU patients across the first 3 days and the patient received no CMM on the fourth day, then the patient-specific pharmacist-to-patient ratio would be: (12+8+14)/3 = 11.3 (ratio: 1:11).

#### eMethods 2. Definition of Ventilator Free Days and Probability of Discharge

We defined 28-day ventilator-free days (VFD) as the number of days from day 0 (first ventilation) to day 28 during which a patient was alive and free of mechanical ventilation. Patients who died before day 28 or remained ventilated through day 28 were assigned VFD = 0. For patients extubated and later re-intubated within 28 days, only days not receiving ventilation counted toward VFD.

Modeling length of stay (LOS) as time to discharge alive with a competing risk of death directly targets the clinical and operational question: “How quickly are patients discharged, accounting for the fact that some die before discharge?” Standard survival or linear LOS models can be misleading because death truncates LOS and creates informative censoring: patients who die are not “long-stayers,” they are non-discharged. The Fine–Gray hazards approach estimates the cumulative incidence of discharge, which (i) treats death appropriately as a competing event rather than censoring it, (ii) yields subdistribution hazard ratios (SHR) that map to an absolute, time-indexed probability of discharge, and (iii) aligns with bed-flow and staffing decisions grounded in probabilities of discharge over time. Practically, SHR >1 indicates a higher rate of discharge (shorter LOS) after adjusting for covariates, while SHR <1 indicates prolonged stays. This framework avoids survivor bias inherent in analyzing raw LOS among only survivors, preserves an interpretable link to absolute risk, and complements hazards models by answering the policy-relevant question about discharge likelihood in the presence of death.

#### eMethods 3. Handling of Missing Data

Missing covariates were addressed under a missing-at-random (MAR) assumption using multiple imputation by chained equations (m = 10). The imputation included all analysis covariates, and the outcomes themselves were not imputed. Continuous variables were imputed with predictive mean matching (k = 5) with logical bounds; binary variables with logistic regression; and multi-category variables with polytomous regression. Derived variables were passively imputed to maintain algebraic consistency. Each analysis model (GEE for mortality, Fine-Gray for LOS) was fit within each imputed dataset, and point estimates and variances were pooled with Rubin’s rules. Convergence diagnostics, distributions of imputed vs observed values, and the fraction of missing information were reviewed and were consistent with the MAR assumption. As sensitivity analyses, we repeated all models in the complete-case cohort and explored pattern-mixture (delta) adjustments to assess robustness to plausible departures from MAR.

### Subgroup analysis - Pediatric ICU

#### Table S1. Patient demographics of pediatric ICU patients

|  | **Overall** | **Alive** | **Deceased** | **p-value** |
| --- | --- | --- | --- | --- |
|  | **N = 786** | **N = 765** | **N = 21** |  |
| ***Primary Variable*** | | | | |
| **CMM with rounding every day of ICU stay** |  |  |  | 0.03 |
| No | 540 (68.7) | 521 (68.1) | 19 (90.5) |  |
| Yes | 246 (31.3) | 244 (31.9) | 2 (9.5) |  |
| ***Secondary Variable*** | | | | |
| **Pharmacist-to-ICU-patient ratio** | 19.0 (14.0, 26.4) | 19.0 (14.0, 26.5) | 19.0 (17.1, 25.3) | 0.70 |
| Missing | 14.0 (1.8%) | 14.0 (1.8%) | 0 (0%) |  |
| ***Co-variates*** | | | | |
| **Age** | 2.9 (0.5, 10.2) | 3.1 (0.5, 10.3) | 0.9 (0, 5.0) | 0.03 |
| **Sex** |  |  |  | >0.9 |
| Male | 418 (53.2) | 407 (53.2) | 11 (52.5) |  |
| Female | 367 (46.7) | 357 (46.7) | 10 (47.6) |  |
| Unidentified | 1 (0.1) | 1 (0.1) | 0 (0) |  |
| **MRC-ICU Score*** | 5.0 (2.0, 11.0) | 5.0 (2.0, 10.0) | 15.0 (11.0, 23.0) | <0.001 |
| **PRISM III score*** | 3.0 (0.0, 7.0) | 3.0 (0.0, 6.0) | 17.0 (10.0, 22.0) | <0.001 |
| Missing | 3 (0.4%) | 3 (0.4%) | 0 (0%) |  |
| **ICU Admission Day of the Week** |  |  |  | 0.30 |
| Monday | 113 (14.4%) | 108 (14.1%) | 5 (23.8%) |  |
| Tuesday | 132 (16.8%) | 130 (17.0%) | 2 (9.5%) |  |
| Wednesday | 131 (16.7%) | 129 (16.9%) | 2 (9.5%) |  |
| Thursday | 112 (14.3%) | 111 (14.5%) | 1 (4.8%) |  |
| Friday | 119 (15.1%) | 116 (15.2%) | 3 (14.3%) |  |
| Saturday | 70 (8.9%) | 68 (8.9%) | 2 (9.5%) |  |
| Sunday | 109 (13.9%) | 103 (13.5%) | 6 (28.6%) |  |
| **Institution Type** |  |  |  |  |
| Academic Medical Center | 786 (100%) | 765 (100%) | 21 (100%) |  |
| Community Hospital | 0 (0.0%) | 0 (0.0%) | 0 (0.0%) |  |
| **ICU pharmacist coverage 1^st^ 24 hours‡** |  |  |  | 0.40 |
| CMM delivered on interprofessional rounds | 547 (69.7%) | 533 (69.8%) | 14 (66.7%) |  |
| CMM delivered not on interprofessional rounds | 97 (12.4%) | 96 (12.6%) | 1 (4.8%) |  |
| Abbreviated CMM delivered | 37 (4.7%) | 36 (4.7%) | 1 (4.8%) |  |
| CMM not delivered (absence of CMM) | 104 (13.3%) | 99 (13.0%) | 5 (23.8%) |  |
| Missing | 1 (0.1%) | 1 (0.1%) | 0 (0.0%) |  |
| **Nurse-to-patient ratio (average), 1:X** |  |  |  | <0.001 |
| ≤1 | 93 (12.1%) | 86 (11.5%) | 7 (33.3%) |  |
| =2 | 460 (59.7%) | 456 (60.9%) | 4 (19.1%) |  |
| >2 | 16 (2.1%) | 16 (2.1%) | 0 (0.0%) |  |
| 1-2 | 201 (26.10%) | 191 (25.50%) | 10 (47.62%) |  |
| Missing | 16 (2.0%) | 16 (2.1%) | 0 (0%) |  |
| **Percentage of medical teams rounded with§** | 50 (50, 72) | 50 (50, 72) | 60 (50, 69) | 0.13 |
| Missing | 19 (2.4) | 19 (2.5) | 0 (0) |  |
| **Renal replacement therapy** |  |  |  | 0.30 |
| No | 773 (98.4%) | 753 (98.4%) | 20 (95.2%) |  |
| Yes | 13 (1.7%) | 12 (1.6%) | 1 (4.8%) |  |
| **Mechanical Circulatory Support** |  |  |  | <0.001 |
| No | 630 (80.2%) | 629 (82.2%) | 1 (4.8%) |  |
| Yes | 156 (19.9%) | 136 (17.8%) | 20 (95.2%) |  |
| **Mechanical ventilation** |  |  |  | <0.001 |
| No | 512 (65.1%) | 511 (66.8%) | 1 (4.8%) |  |
| Yes | 274 (34.9%) | 254 (33.2%) | 20 (95.2%) |  |
| ***Outcomes*** | | | | |
| **Hospital Length of Stay** | 4.24 (2.5, 8.8) | 4.12 (2.4, 8.6) | 11.80 (5.2, 48.5) | <0.001 |
| **ICU Length of Stay** | 2.7 (1.0, 5.3) | 2.5 (1.0, 5.0) | 11.0 (5.0, 39.0) | <0.001 |
| **ICU Free Days** | 25.4 (22.7, 27.0) | 25.5 (23.0, 27.0) | 0.0 (0.0, 0.0) | <0.001 |
| Missing | 8 (1.0%) | 1 (0.1%) | 7 (33.0%) |  |
| **Duration of mechanical ventilation** | 0 (0, 0.7) | 0 (0, 0.4) | 6.0 (3.5, 9.0) | <0.001 |
| **Ventilator Free Days** | 28.0 (27.3, 28.0) | 28.0 (27.6, 28.0) | 0 (0, 0) | <0.001 |

CMM: comprehensive Medication Management; ICU: Intensive Care Unit; MRC-ICU: medication regimen complexity-intensive care unit; PRISM III: Pediatric Risk of Mortality III

Data reported as n (%) or median (interquartile range)

**‡**CMM delivered on interprofessional rounds: the pharmacist attended multidisciplinary rounds and verbally provided CMM (including review of medications and recommendations); CMM delivered not on interprofessional rounds: the pharmacist provided by CMM (reviewed medications and provided recommendations) but did not attend interdisciplinary rounds; Abbreviated CMM delivered: the pharmacist provided abbreviated CMM which includes brief medication review but does not include full patient review (e.g. progress notes, results) and recommendations were provided outside of rounds; CMM not delivered (absence of CMM): the patient received no medication review from a pharmacist outside of pharmacokinetic monitoring and prospective medication order verification

*worst score during the first 24 hours of ICU stay

**§**Percent of assigned ICU teams pharmacist rounded is calculated by total number of ICU medical teams the pharmacist attended and provided CMM on interprofessional rounds for divided by total number of ICU medical teams providing care for patients that the pharmacist was assigned to provide CMM for

#### Table S2. Regression Analyses for Mortality and Fine-Gray Competing Risks Model for Length of Stay and Ventilator Free Day with In-ICU Mortality as a Competing Event in Pediatric ICU Patients

| **Mortality** | | | | | **Hazard of Discharge Alive from ICU** | | **Hazard of Discharge Alive from Hospital** | | **Hazard of Extubation Alive** | |
| --- | --- | --- | --- | --- | --- | --- | --- | --- | --- | --- |
| **Variable** | **Univariable** | | **Multivariable** | |  |  |  |  |  |  |
|  | **OR (CI)** | **P-value** | **OR (CI)** | **P-value** | **Hazard Ratio (CI)** | **P-value** | **Hazard Ratio (CI)** | **P-value** | **Hazard Ratio (CI)** | **P-value** |
| ***Primary Variable*** | | | | | | | | | | |
| **CMM with rounding every day of ICU stay** | 0.27 (0.07, 1.03) | 0.06 | 0.15 (0.03, 0.79) | 0.03 | 2.29 (1.91-2.74) | <0.001 | 1.99 (1.61–2.46) | < 0.001 | 1.38 (1.23–1.55) | < 0.001 |
| ***Secondary Variable*** | | | | | | | | | | |
| **Pharmacist-to-ICU-patient ratio** | 0.99 (0.95, 1.04) | 0.75 | 1.06 (1.00, 1.14) | 0.11 | 0.99 (0.98-1.00) | 0.03 | 1.00 (1.00–1.01) | 0.80 | 0.99 (0.99–1.00) | < 0.001 |
| ***Co-variates*** | | | | | | | | | | |
| **Age** | 0.93 (0.85, 1.01) | 0.10 | 0.93 (0.85, 1.02) | 0.12 | 1.02 (1.01-1.04) | <0.001 | 1.02 (1.00–1.03) | 0.029 | 1.03 (1.02–1.04) | < 0.001 |
| **Sex: Male§** | - | - | - | - | - | - | - | - | - | - |
| Sex: Female | 1.04 (0.45, 2.43) | 0.93 | 1.08 (0.43, 2.70) | 0.88 | 0.98 (0.86-1.13) | 0.79 | 1.00 (0.86–1.17) | 0.97 | 1.09 (0.98–1.20) | 0.11 |
| **MRC-ICU Score** | 1.15 (1.09, 1.20) | <0.001 | 1.05 (1.00, 1.11) | 0.10 | 0.97 (0.96-0.98) | <0.001 | 0.97 (0.96–0.98) | < 0.001 | 0.95 (0.94–0.96) | < 0.001 |
| **PRISM III Score** | 1.25 (1.17, 1.32) | <0.001 | 1.19 (1.11, 1.27) | <0.001 | 0.96 (0.94-0.98( | <0.001 | 0.95 (0.94–0.97) | < 0.001 | 0.95 (0.94–0.97) | < 0.001 |
| **ICU Admission Day of the Week: Monday§** | - | - | - | - | - | - | - | - | - | - |
| ICU Admission Day of the Week: Tuesday | 0.38 (0.08, 1.72) | 0.21 | 0.26 (0.04,1.60) | 0.15 | 1.10 (0.88-1.38) | 0.42 | 1.11 (0.83–1.48) | 0.48 | 1.12 (0.94–1.34) | 0.22 |
| ICU Admission Day of the Week: Wednesday | 0.38 (0.08, 1.74) | 0.21 | 0.56 (0.11, 3.00) | 0.50 | 1.19 (0.95-1.49) | 0.12 | 1.15 (0.88–1.50) | 0.32 | 1.08 (0.89–1.32) | 0.44 |
| ICU Admission Day of the Week: Thursday | 0.27 (0.04, 1.65) | 0.15 | 0.24 (0.04, 1.60) | 0.14 | 1.17 (0.92-1.50) | 0.21 | 1.13 (0.86–1.50) | 0.38 | 1.17 (0.98–1.40) | 0.08 |
| ICU Admission Day of the Week: Friday | 0.59 (0.15, 2.32) | 0.45 | 0.55 (0.10, 3.10) | 0.50 | 1.01 (0.76-1.32) | 0.97 | 0.94 (0.67–1.31) | 0.72 | 1.25 (1.02–1.53) | 0.04 |
| ICU Admission Day of the Week: Saturday | 0.72 (0.16, 3.31) | 0.67 | 0.61 (0.08, 4.53) | 0.63 | 1.17 (0.77-1.76) | 0.47 | 1.06 (0.70–1.62) | 0.79 | 1.10 (0.80–1.52) | 0.57 |
| ICU Admission Day of the Week: Sunday | 1.24 (0.39, 4.00) | 0.72 | 2.45 (0.55, 10.94) | 0.72 | 0.79 (0.61-1.04) | 0.09 | 0.79 (0.59–1.07) | 0.13 | 0.89 (0.71–1.11) | 0.30 |
| **1st 24h of ICU: Pharmacist rounded with team§** | - | - | - | - | - | - | - | - | - | - |
| 1st 24h of ICU: Pharmacist without rounding | 0.57 (0.11, 3.11) | 0.52 | 1.04]1 (0.20, 5.03) | 0.99 | 1.21 (0.96-1.53) | 0.10 | 1.20 (0.95–1.52) | 0.05 | 1.00 (0.85–1.20) | 0.92 |
| 1st 24h of ICU: Profile review only | 1.50 (0.27, 8.35) | 0.64 | 0.67 (0.08, 5.77) | 0.71 | 1.30 (0.88-1.91) | 0.18 | 0.83 (0.58–1.21) | 0.34 | 1.06 (0.84–1.35) | 0.62 |
| 1st 24h of ICU: Not reviewed | 2.03 (0.74, 5.54) | 0.17 | 1.97 (0.49, 7.94) | 0.34 | 1.10 (0.82-1.47) | 0.52 | 1.14 (0.84–1.53) | 0.40 | 0.88 (0.70–1.10) | 0.29 |
| **RN-to-ICU-patient ratio: =2§** | - | - | - | - | - | - | - | - | - | - |
| RN-to-ICU-patient ratio: <=1 | 8.88 (2.70, 29.21) | <0.001 | 5.53 (1.23, 24.80) | 0.03 | 1.22 (0.9301.61) | 0.15 | 0.85 (0.63–1.15) | 0.30 | 0.92 (0.79–1.08) | 0.33 |
| RN-to-ICU-patient ratio: 1-2 | 5.66 (1.85, 17.30) | 0.002 | 3.70 (1.12, 12.20) | 0.03 | 0.60 (0.51-0.72) | <0.001 | 0.54 (0.45–0.65) | < 0.001 | 0.79 (0.68–0.92) | 0.003 |
| RN-to-ICU-patient ratio: >2 | 2.90 (0.15, 56.22) | 0.48 | 19.59 (1.32, 290) | 0.03 | 0.57 (0.37-0.86) | 0.01 | 0.67 (0.44–1.02) | 0.06 | 1.00 (0.83–1.17) | 0.88 |
| **Percentage of medical teams rounded with** | 1.72 (0.28, 10.74) | 0.56 | 37.22 (1.96, 707) | 0.02 | 0.74 (0.50-1.10) | 0.13 | 0.79 (0.49–1.27) | 0.33 | 0.70 (0.55–0.88) | 0.002 |

CMM: comprehensive Medication Management; ICU: Intensive Care Unit; MRC-ICU: medication regimen complexity-intensive care unit; PRISM III: Pediatric Risk of Mortality III

*worst score during the first 24 hours of ICU stay

**‡**CMM delivered on interprofessional rounds: the pharmacist attended multidisciplinary rounds and verbally provided CMM (including review of medications and recommendations); CMM delivered not on interprofessional rounds: the pharmacist provided by CMM (reviewed medications and provided recommendations) but did not attend interdisciplinary rounds; Abbreviated CMM delivered: the pharmacist provided abbreviated CMM which includes brief medication review but does not include full patient review (e.g. progress notes, results) and recommendations were provided outside of rounds; CMM not delivered (absence of CMM): the patient received no medication review from a pharmacist outside of pharmacokinetic monitoring and prospective medication order verification

**§**Percent of assigned ICU teams pharmacist rounded is calculated by total number of ICU medical teams the pharmacist attended and provided CMM on interprofessional rounds for divided by total number of ICU medical teams providing care for patients that the pharmacist was assigned to provide CMM for

### Subgroup analysis - Neonatal ICU

#### Table S3. Patient demographics for neonatal ICU patients

|  | **Overall** | **Alive** | **Deceased** | **p-value** |
| --- | --- | --- | --- | --- |
|  | **N = 563** | **N = 547** | **N = 16** |  |
| ***Primary Variable*** | | | | |
| **CMM with rounding every day of ICU stay‡** |  |  |  | 0.40 |
| No | 513 (91.1%) | 497 (90.9%) | 16 (100%) |  |
| Yes | 50 (8.9%) | 50 (9.1%) | 0 (0.00%) |  |
| ***Secondary Variable*** | | | | |
| **Pharmacist-to-ICU-patient ratio** | 38.2 (30.5, 44.0) | 38.2 (30.5, 44.0) | 38.5 (29.6, 44.8) | 0.80 |
| Missing | 4.0 (0.7%) | 4.0 (0.7%) | 0.0 (0%) |  |
| ***Co-variates*** | | | | |
| **Age, years** | 0.0 (0.0, 0.0) | 0.0 (0.0, 0.0) | 0.0 (0.0, 0.0) | 0.10 |
| **Sex** |  |  |  | >0.9 |
| Male | 302 (53.6%) | 293 (53.6%) | 9 (56.3%) |  |
| Female | 260 (46.2%) | 253 (46.3%) | 7 (43.8%) |  |
| Unidentified | 1 (0.2%) | 1 (0.2%) | 0 (0.0%) |  |
| **MRC-ICU Score*** | 4.0 (1.0, 7.0) | 4.0 (1.0, 7.0) | 9.5 (7.5, 13.5) | <0.001 |
| **PRISM III score*** | 3.0 (0.0, 8.0) | 3.0 (0.0, 8.0) | 12.0 (9.0, 15.0) | <0.001 |
| Missing | 2 (0.4%) | 2 (0.4%) | 0 (0.0%) |  |
| **ICU Admission Day of the Week** |  |  |  | 0.50 |
| Monday | 69 (12.3%) | 65 (11.9%) | 4 (25.0%) |  |
| Tuesday | 111 (19.7%) | 109 (19.9%) | 2 (12.5%) |  |
| Wednesday | 90 (16.0%) | 88 (16.1%) | 2 (12.5%) |  |
| Thursday | 90 (16.0%) | 89 (16.3%) | 1 (6.3%) |  |
| Friday | 86 (15.3%) | 82 (15.0%) | 4 (25.0%) |  |
| Saturday | 55 (9.8%) | 53 (9.7%) | 2 (12.5%) |  |
| Sunday | 62 (11.0%) | 61 (11.2%) | 1 (6.3%) |  |
| **Institution Type** |  |  |  | >0.9 |
| Academic Medical Center | 352 (62.5%) | 342 (62.5%) | 10 (62.5%) |  |
| Community Hospital | 211 (37.5%) | 205 (37.5%) | 6 (37.5%) |  |
| **ICU pharmacist coverage 1^st^ 24 hours‡** |  |  |  | 0.8 |
| CMM delivered on interprofessional rounds | 348 (61.9%) | 338 (61.9%) | 10 (62.5%) |  |
| CMM delivered not on interprofessional rounds | 79 (14.1%) | 77 (14.1%) | 2 (12.5%) |  |
| Abbreviated CMM delivered | 22 (3.9%) | 21 (3.9%) | 1 (6.3%) |  |
| CMM not delivered (absence of CMM) | 113 (20.1%) | 110 (20.2%) | 3 (18.8%) |  |
| Missing | 1 (0.2%) | 1 (0.2%) | 0 (0%) |  |
| **Nurse-to-patient ratio (average), 1:X** |  |  |  | <0.001 |
| ≤1 | 4 (0.7%) | 3 (0.6%) | 1 (6.3%) |  |
| =2 | 231 (41.5%) | 227 (42.0%) | 4 (25.0%) |  |
| >2 | 233 (41.8%) | 231 (42.7%) | 2 (12.5%) |  |
| 1-2 | 89 (16.0%) | 80 (15.0%) | 9 (56.3%) |  |
| Missing | 6 (1.1%) | 6 (1.1%) | 0 (0%) |  |
| **Percentage of medical teams rounded with§** | 0.69 (0.6, 1.0) | 0.69 (0.6, 1.0) | 0.62 (0.5, 1.0) | 0.40 |
| Missing | 4 (0.7%) | 4 (0.7%) | 0 (0.0%) |  |
| **Renal replacement therapy** |  |  |  | >0.90 |
| No | 563 (100%) | 547 (100%) | 16 (100%) |  |
| Yes | 0 (0.0%) | 0 (0.0%) | 0 (0.0%) |  |
| **Mechanical Circulatory Support** |  |  |  | <0.001 |
| No | 524 (93.1%) | 519 (94.9%) | 5 (31.3%) |  |
| Yes | 39 (6.9%) | 28 (5.1%) | 11 (68.8%) |  |
| **Mechanical ventilation** |  |  |  | <0.001 |
| No | 407 (72.3%) | 406 (74.2%) | 1 (6.3%) |  |
| Yes | 156 (27.7%) | 141 (25.7%) | 15 (93.8%) |  |
| ***Outcomes*** | | | | |
| **Hospital Length of Stay, days** | 12.6 (5.5, 37.2) | 12.60 (5.8, 37.2) | 17.8 (4.7, 39.1) | >0.90 |
| **ICU Length of Stay, days** | 12.0 (5.0, 35.0) | 12.0 (5.0, 35.0) | 17.0 (4.5, 39.0) | 0.80 |
| **ICU Free Days** | 16.0 (0.0, 23.0) | 16.0 (0.0, 23.0) | 0.0 (0.0, 0.0) | <0.001 |
| Missing | 10 (1.8%) | 6 (1.1%) | 4 (25.0%) |  |
| **Duration of mechanical ventilation, days** | 0.0 (0.0, 0.1) | 0.0 (0.0, 0.00) | 4.1 (0.9, 19.6) | <0.001 |
| **Ventilator Free Days** | 28.0 (27.9, 28.0) | 28.0 (28.0, 28.0) | 0.0 (0.0, 0.0) | <0.001 |

CMM: comprehensive Medication Management; ICU: Intensive Care Unit; MRC-ICU: medication regimen complexity-intensive care unit; PRISM III: Pediatric Risk of Mortality III

Data reported as n (%) or median (interquartile range)

**‡**CMM delivered on interprofessional rounds: the pharmacist attended multidisciplinary rounds and verbally provided CMM (including review of medications and recommendations); CMM delivered not on interprofessional rounds: the pharmacist provided by CMM (reviewed medications and provided recommendations) but did not attend interdisciplinary rounds; Abbreviated CMM delivered: the pharmacist provided abbreviated CMM which includes brief medication review but does not include full patient review (e.g. progress notes, results) and recommendations were provided outside of rounds; CMM not delivered (absence of CMM): the patient received no medication review from a pharmacist outside of pharmacokinetic monitoring and prospective medication order verification

*worst score during the first 24 hours of ICU stay

**§**Percent of assigned ICU teams pharmacist rounded is calculated by total number of ICU medical teams the pharmacist attended and provided CMM on interprofessional rounds for divided by total number of ICU medical teams providing care for patients that the pharmacist was assigned to provide CMM for

#### Table S4. Regression Analyses for Mortality and Fine-Gray Competing Risks Model for Length of Stay and Ventilator Free Day with In-ICU Mortality as a Competing Event in Neonatal ICU Patients

| **Variable** | | **Mortality** | | | | **Hazard of Discharge Alive from ICU** | | **Hazard of Discharge Alive from Hospital** | | **Hazard of Extubation Alive** | |
| --- | --- | --- | --- | --- | --- | --- | --- | --- | --- | --- | --- |
|  |  | **Univariable** | | **Multivariable** | |  |  |  |  |  |  |
|  |  | **OR (CI)** | **P-value** | **OR (CI)** | **P-value** | **Hazard Ratio (CI)** | **P-value** | **Hazard Ratio (CI)** | **P-value** | **Hazard Ratio (CI)** | **P-value** |
| ***Primary Variable*** | | | | | | | | | | | |
| **CMM with rounding every day of ICU stay** | | 0.30 (0.02, 5.05) | 0.40 | 1.08 (0.09, 13.00) | 0.95 | 5.78 (3.92–8.53) | <0.001 | 5.67 (3.41–9.42) | < 0.001 | 0.98 (0.84–1.14) | 0.77 |
| ***Secondary Variable*** | | | | | | | | | | | |
| **Pharmacist-to-ICU-patient ratio** | | 1.00 (0.94, 1.06) | 0.93 | 1.01 (0.93, 1.10) | 0.75 | 1.03 (1.01–1.04) | 0.01 | 1.02 (1.00–1.04) | 0.09 | 1.00 (0.99–1.01) | 0.46 |
| **Co-variates** | | | | | | | | | | | |
| **Sex: Male§** | **-** | | - | - | - | - | - | - | - | - | - |
| Sex, female | | 0.91 (0.35, 2.42) | 0.86 | 0.71 (0.28, 1.81) | 0.47 | 0.77 (0.66–0.91) | 0.001 | 0.82 (0.68–0.98) | 0.031 | 0.96 (0.87–1.07) | 0.50 |
| **MRC-ICU Score*** | | 1.20 (1.11, 1.30) | <0.001 | 1.08 (0.99, 1.19) | 0.10 | 0.93 (0.91–0.951) | <0.001 | 0.92 (0.89–0.94) | < 0.001 | 0.96 (0.94–0.97) | < 0.001 |
| **PRISM III*** | | 1.16 (1.09, 1.24) | <0.001 | 1.06 (0.99, 1.14) | 0.110 | 0.92 (0.90–0.94) | <0.001 | 0.92 (0.90–0.93) | < 0.001 | 0.96 (0.94–0.97) | < 0.001 |
| **ICU admission day of the week: Monday** | | - | - | - | - | - | - | - | - | - | - |
| ICU Admission Day of the Week: Tuesday | | 0.33 (0.07, 1.61) | 0.17 | 0.53 (0.11,2.51) | 0.42 | 1.19 (0.84–1.68) | 0.33 | 1.32 (0.93–1.87) | 0.12 | 1.13 (0.90–1.43) | 0.29 |
| ICU Admission Day of the Week: Wednesday | | 0.41 (0.09, 1.99) | 0.27 | 0.28 (0.05, 1.50) | 0.14 | 1.65 (1.17–2.33) | 0.01 | 1.64 (1.15–2.34) | 0.01 | 1.18 (0.94–1.47) | 0.16 |
| ICU Admission Day of the Week: Thursday | | 0.24 (0.04, 1.59) | 0.140 | 0.33 (0.06, 2.02) | 0.23 | 1.19 (0.83–1.71) | 0.35 | 1.34 (0.92–1.95) | 0.13 | 1.14 (0.90–1.44) | 0.29 |
| ICU Admission Day of the Week: Friday | | 0.79 (0.21, 3.05) | 0.74 | 0.98 (0.22, 4.40) | 0.98 | 1.51 (1.01–2.28) | 0.05 | 1.25 (0.77–2.01) | 0.37 | 0.99 (0.73–1.34) | 0.93 |
| ICU Admission Day of the Week: Saturday | | 0.68 (0.14, 3.33) | 0.63 | 0.679 (0.11, 5.85) | 0.81 | 1.93 (1.15–3.24) | 0.01 | 1.43 (0.78–2.59) | 0.25 | 1.22 (0.84–1.77) | 0.31 |
| ICU Admission Day of the Week: Sunday | | 0.36 (0.05, 2.33) | 0.28 | 0.42 (0.06, 3.13) | 0.39 | 1.20 (0.79–1.80) | 0.40 | 1.28 (0.85–1.94) | 0.24 | 1.15 (0.90–1.48) | 0.26 |
| **Hospital type (reference: academic medical center)** | | - | - | - | - | - | - | - | - | - | - |
| Community Hospital | | 1.03 (0.38, 2.79) | 0.95 | 45.68 (2.51, 832) | 0.01 | 2.58 (1.30–5.13) | 0.01 | 2.05 (1.03–4.09) | 0.04 | 0.88 (0.66–1.18) | 0.40 |
| **1st 24 hours of ICU: Pharmacist rounded with team‡ (reference: CMM delivered on interprofessional rounds)** | | - | - | - | - | - | - | - | - | - | - |
| CMM delivered not on interprofessional rounds | | 1.04 (0.26, 4.22) | 0.96 | 1.46 (0.31, 6.95) | 0.63 | 1.26 (0.97–1.63) | 0.09 | 1.20 (0.87–1.67) | 0.27 | 1.06 (0.89–1.26) | 0.55 |
| Abbreviated CMM delivered | | 2.24 (0.38, 13.11) | 0.37 | 0.76 (0.08, 7.16) | 0.81 | 0.44 (0.20–0.97) | 0.04 | 0.55 (0.26–1.16) | 0.12 | 0.78 (0.51–1.20) | 0.26 |
| CMM not delivered (absence of CMM) | | 1.02 (0.30, 3.50) | 0.97 | 0.81 (0.18, 3.64) | 0.79 | 0.70 (0.51–0.95) | 0.02 | 0.90 (0.64–1.26) | 0.53 | 0.99 (0.79–1.23) | 0.90 |
| **Nurse-to-patient ratio (average) (reference: 1:2)** | | - | - | - | - | - | - | - | - | - | - |
| ≤1 | | 20.43 (2.44, 171) | 0.01 | 11.91 (1.00, 142) | 0.05 | 6.11 (3.14–11.91) | < 0.001 | 2.32 (0.65–8.27) | 0.20 | 0.64 (0.30–1.35) | 0.24 |
| 1-2 | | 6.00 (1.90, 18.95) | 0.002 | 3.53 (1.07, 11.70) | 0.04 | 0.49 (0.36–0.66) | <0.001 | 0.57 (0.41–0.77) | < 0.001 | 0.68 (0.54–0.85) | < 0.001 |
| >2 | | 0.55 (0.12, 2.62) | 0.46 | 0.13 (0.03, 0.63) | 0.01 | 0.37 (0.26–0.54) | <0.001 | 0.55 (0.37–0.81) | 0.003 | 1.14 (0.93–1.39) | 0.20 |
| **Percent of assigned ICU teams pharmacist rounded with§** | | 0.38 (0.04, 3.34) | 0.38 | 0.01 (0.00, 1.28) | 0.06 | 0.27 (0.08–0.92) | 0.04 | 0.24 (0.07–0.89) | 0.03 | 1.01 (0.64–1.58) | 0.97 |

CCP: Critical Care Pharmacist; CMM: comprehensive Medication Management; ICU: Intensive Care Unit; MRC-ICU: medication regimen complexity-intensive care unit; PRISM III: Pediatric Risk of Mortality III

*worst score during the first 24 hours of ICU stay

**‡**CMM delivered on interprofessional rounds: the pharmacist attended multidisciplinary rounds and verbally provided CMM (including review of medications and recommendations); CMM delivered not on interprofessional rounds: the pharmacist provided by CMM (reviewed medications and provided recommendations) but did not attend interdisciplinary rounds; Abbreviated CMM delivered: the pharmacist provided abbreviated CMM which includes brief medication review but does not include full patient review (e.g. progress notes, results) and recommendations were provided outside of rounds; CMM not delivered (absence of CMM): the patient received no medication review from a pharmacist outside of pharmacokinetic monitoring and prospective medication order verification

**§**Percent of assigned ICU teams pharmacist rounded is calculated by total number of ICU medical teams the pharmacist attended and provided CMM on interprofessional rounds for divided by total number of ICU medical teams providing care for patients that the pharmacist was assigned to provide CMM for
